# Loss-of-function of the Zinc Finger Homeobox 4 (*ZFHX4*) gene underlies a neurodevelopmental disorder

**DOI:** 10.1101/2024.08.07.24311381

**Authors:** María del Rocío Pérez Baca, María Palomares Bralo, Michiel Vanhooydonck, Lisa Hamerlinck, Eva D’haene, Sebastian Leimbacher, Eva Z. Jacobs, Laurenz De Cock, Erika D’haenens, Annelies Dheedene, Zoë Malfait, Lies Vantomme, Ananilia Silva, Kathleen Rooney, Fernando Santos-Simarro, Roser Lleuger-Pujol, Sixto García-Miñaúr, Itsaso Losantos-García, Björn Menten, Gaia Gestri, Nicola Ragge, ZFHX4 consortium, Bekim Sadikovic, Elke Bogaert, Delfien Syx, Bert Callewaert, Sarah Vergult

## Abstract

8q21.11 microdeletions encompassing the gene encoding transcription factor ZFHX4, have previously been associated by us with a syndromic form of intellectual disability, hypotonia, decreased balance and hearing loss. Here, we report on 57 individuals, 52 probands and 5 affected family members, with protein truncating variants (n=36), (micro)deletions (n=20) or an inversion (n=1) affecting *ZFHX4* with variable developmental delay and intellectual disability, distinctive facial characteristics, morphological abnormalities of the central nervous system, behavioral alterations, short stature, hypotonia, and occasionally cleft palate and anterior segment dysgenesis. The phenotypes associated with 8q21.11 microdeletions and *ZFHX4* intragenic loss-of-function variants largely overlap, identifying ZFHX4 as the main driver for the microdeletion syndrome, although leukocyte-derived DNA shows a mild common methylation profile for (micro)deletions only. We identify ZFHX4 as a transcription factor that is increasingly expressed during human brain development and neuronal differentiation. Furthermore, ZFHX4 interacting factors identified via IP-MS in neural progenitor cells, suggest an important role for ZFHX4 in cellular and developmental pathways, especially during histone modifications, cytosolic transport and development. Additionally, using CUT&RUN, we observed that ZFHX4 binds with the promoter regions of genes with crucial roles in embryonic, neuron and axon development. Since loss-of-function variants in *ZFHX4* are found with consistent dysmorphic facial features, we investigated whether the disruption of *zfhx4* causes craniofacial abnormalities in zebrafish. First-generation (F0) *zfhx4* crispant zebrafish, (mosaic) mutant for *zfhx4* loss-of-function variants, have significantly shorter Meckel’s cartilages and smaller ethmoid plates compared to control zebrafish. Furthermore, behavioral assays show a decreased movement frequency in the *zfhx4* crispant zebrafish in comparison with control zebrafish larvae. Although further research is needed, our *in vivo* work suggests a role for zfhx4 in facial skeleton patterning, palatal development and behavior.

## Introduction

Neurodevelopmental disorders (NDDs) affect approximately 2 to 5% of the children worldwide. In up to 40% of the affected individuals, a monogenic cause can be identified, but many gene-disease relationships in NDDs remain to be discovered^1^. In 2011, Palomares *et al.* reported eight unrelated individuals with a recognizable intellectual disability (ID) syndrome, all sharing a (sub)microscopic deletion in the 8q21.11 region containing *ZFHX4*^2^. Since then, others reported individuals with 8q21.11 deletions ranging from 0.66 to 14.5 Mb in size, all with similar clinical characteristics^3–7^. In 2019, a loss-of-function *ZFHX4* splice variant was reported in a 7-year-old male from a cohort of patients with apraxia of speech^8^. In 2020, *ZFHX4* was identified as a novel NDD candidate gene in a large-scale exome-sequencing study^9^. Hence, *ZFHX4* haploinsufficiency might be one of the drivers of the 8q21.11 microdeletion phenotype. In addition, Bishop *et al.* identified *ZFHX4* as a novel orofacial cleft candidate gene^10^. In both exome driven studies, *de novo ZFHX4* loss-of-function (LoF) variants appeared to be enriched in the affected individuals. In alignment with these data, the latest gnomAD version (v4.1.0) predicts *ZFHX4* to be intolerant to protein truncating variants (PTVs), with a pLI score (probability of being loss-of-function intolerant) of 1, and a constraint score of observed/expected of 0.24^11^.

*ZFHX4* comprises 12 exons and encodes a transcription factor that carries 22 zinc fingers and four homeodomains^12^. Expression of ZFHX4 is high in the developing brain of different species^1,3–5^, and shown to decrease in adult mice ^13^. In addition, Zfhx4 is highly expressed in cartilage^17^. In mice, Zfhx4 deficiency is associated with cleft palate, and its expression in the palatal shelves supports a role for Zfhx4 in palatal development^17^. Homozygous *Zfhx4* knockout mice die of neurogenic respiratory disorders within 24 hours after birth^18^. In humans, ZFHX4 expression has been detected in adult brain, muscle, and liver^12^. Altogether, ZFHX4 has been identified as a transcription factor involved in neural cell maturation, region-specific brain differentiation^14^ and skeletal development^17^.

Moreover, in accordance with a role in cell maturation, ZFHX4 appears to be involved in the biology of different types of cancer, such as glioblastoma^19^, esophageal squamous cell carcinoma (ESCC)^20,21^ and ovarian cancer^22,23^. Chudnovsky *et al.* showed that ZFHX4 regulates therapy-resistant tumour initiating (TIC) glioblastoma cell state by interacting with CHD4, a core member of the nucleosome remodeling and deacetylase (NuRD) complex^19^. Additionally, several studies showed that in ESCC and ovarian cancer, ZFHX4 expression can be used as a prognostic biomarker for survival outcomes in patients with metastasis^22^.

Here, we report a cohort of 57 individuals with *ZFHX4* aberrations. We evaluate the common features and differences between *ZFHX4* protein truncating variants (PTVs) and copy number variants (CNVs) including *ZFHX4* to inform clinical management. Our observations confirm that ZFHX4 loss-of-function drives the microdeletion phenotype. Furthermore, we evaluated the protein and DNA interactome of ZFHX4, indicating an important role for ZFHX4 in normal cell and tissue maturation, and in neurodevelopmental pathways including axon development. Finally, zebrafish *zfhx4* crispants show craniofacial malformations and behavioral abnormalities. Overall, our data indicate that ZFHX4 is essential for neural and craniofacial development.

## Material and methods

### Patient ascertainment

Individuals with *ZFHX4* alterations were identified through Clinical Genetics Services worldwide using web-based databases such as GeneMatcher^24^ and DECIPHER. The clinical research activity relating to this report has been in accordance with the Declaration of Helsinki on the Ethical Principles for Medical Research Involving Human Subjects. Genetic tests were performed clinically or using research protocols approved by each center ethics committee. Informed consent for genetic analyses, clinical data and biological sample collection, and publication of relevant findings were obtained for all individuals. We collected the detailed phenotype data through a standard form specific for *ZFHX4* (Table S1 and S2).

### Statistics

Statistical studies were performed in SAS 9.4 (SAS Institute, Cary, NC, USA). Comparative studies between females and males and between CNV and LoF variants were calculated using two-tailed Fischer’s exact test. P values <0.05 were considered statistically significant. Counts for clinical characteristics were given only for those where the respective information was available, therefore the total count can be lower than the total number of individuals.

### Facial feature analysis

In order to make the description of facial features as homogeneous as possible, the facial features of all individuals were evaluated by two experienced dysmorphologists, Fernando Santos-Simarro and Sixto García-Miñaúr. The DeepGestalt technology by Face2 Gene (Face2Gene, FDNA Inc., Boston, MA) was used to validate the presence of distinct facial patterns. The cohort was divided into two groups to analyze facial features. The first group included a total of 31 frontal photographs of 20 individuals with CNVs. The second group included 26 facial photographs of 18 individuals with predicted LoF variants in *ZFHX4*. In both cases, age-, sex- and ethnicity-matched healthy individuals were used as a comparison cohort to generate a composite image of the affected individuals. The composite image (not included as a figure) was generated in both cases using DeepGestalt facial analysis and an assessment of the binary comparison between controls and affected individuals was performed by measuring the receiver operating characteristic (ROC) curves and the corresponding area under the curve (AUC) as described by Mak et al. 2021^25^.

### Cell culture

#### Fibroblast culture, treatment and RNA extraction

Fibroblasts derived from skin biopsies were cultured in DMEM medium supplemented with Fetal Calf Serum (FCS), Penicillin/Streptomycin, Kanamycin, Non-Essential Amino Acids (NEAA) and Fungizone until we had at least 600 000 cells. Afterwards, 300,000 cells were seeded in a P60 plate on Day 1 for cycloheximide treatment. On Day 2 the culture medium was replenished with or without cycloheximide (5μg/mL, C7698, Sigma Aldrich) and on Day 3 RNA was extracted with the Maxwell. In short, the cells were loosened with a cell scraper and transferred to a 15mL tube, which was then centrifuged for 3 minutes at 300rcf. Afterwards the supernatant was removed and 200μL 1-thioglycerol/homogenization solution (Promega) was added. The cells were then vortexed and 200μL lysis buffer (Promega) was added. Finally, the sample (400μL) was transferred to a well from the Maxwell and RNA was extracted with the RSC Simply RNA Tissue kit (Promega) following the RSC Simply RNA Cells method.

#### Human embryonic stem cells (hESCs)

The naïve hESCs were cultured according to the protocol described in Pérez Baca et al, 2024^26^.

#### Induced pluripotent cells (hIPSCs)

The iPSC line was commercially obtained from Axol BioScience and maintained in mTESR^TM^ Plus (StemcellTechnologies, #100-0276) on coated cultured plates with Geltrex® LDEV-Free hESC-Qualified Reduced Growth Factor Basement Membrane Matrix (GibcoTM, ThermoFisher), according to the manufacturer’s instructions. The induced pluripotent stem cells were cultured at 37°C in a 5% CO_2_ humid atmosphere. Double volume was replaced every two days as a fixed feeding interval.

#### Neural progenitor cells (NPCs)

The NPCs were differentiated starting with the above mentioned hIPSCs 0.5 x 10^6^ on coated cultured plates with Geltrex® LDEV-Free hESC-Qualified Reduced Growth Factor Basement Membrane Matrix (GibcoTM, ThermoFisher), proceeding with the monolayer protocol using the STEMdiff^TM^ SMADi neural induction kit (StemcellTechnologies, #05835) according to the manufacturer’s instructions for 6-9 days and further cultured up to 18-21 days prior to freezing. The NPCs were cultured at 37°C in a 5% CO_2_ humid atmosphere. Daily full-medium change as a fixed feeding interval until ready to be passaged.

### Hi-C library preparation and analysis

A Hi-C library was generated according to the *in situ* Hi-C protocol adopted by the 4D Nucleome consortium^27^ with a few adaptations. Briefly, five million patient fibroblasts were crosslinked using 2% formaldehyde, lysed, snap frozen in liquid nitrogen and stored at -80°C until further processing. Pre-lysed, crosslinked nuclei were then digested overnight using 250 U *Dpn*II restriction enzyme (New England Biolabs, R0543L). DNA ends were marked by incorporating biotin-14-dATP (Life Technologies, 19524-016) and ligated for 4 hours using 2000 U T4 DNA ligase (New England Biolabs, M0202L). Subsequently, crosslinks were reversed overnight using proteinase K (Qiagen, 19131) and Hi-C template DNA was purified using 1x AMPure XP beads (Beckman Coulter, A63881) and stored at 4°C until library preparation. Hi-C template DNA was sheared to a size of 300-500 bp using microTUBE snap-caps (Covaris, 520045) in a Covaris M220 sonicator and MyOne Streptavidin T1 beads (Life Technologies, 65601) were used to pull down biotinylated ligation junctions. Next, the sample was split into 5 µg aliquots for sequencing library preparation using the NEBNext Ultra II DNA Library Prep Kit (New England Biolabs, E7645L) and NEBNext Multiplex Oligos (New England Biolabs, E7335L). The amplified library was purified, and size selected using 0.55x and 1.2x AMPure XP beads (Beckman Coulter, A63881). Pooled libraries were sequenced on an Illumina NovaSeq 6000 using 100 bp paired-end reads to a depth of ∼500 million reads.

FASTQ files containing raw sequencing data were processed into a Hi-C contact matrix containing both raw and normalized counts using the Juicer pipeline (v1.6)^28^ with BWA-MEM mapping (v0.7.17)^29^ Click or tap here to enter text.to the hg38 reference genome. A processed and normalized Hi-C matrix for control fibroblasts was obtained from Melo *et al.*^30^. We used FAN-C^31^ Click or tap here to enter text.to visualize Knight-Ruiz (KR) normalized and observed/expected patient vs. control Hi-C matrices for the region of interest.

### Expression profiling

RNA of cultured cells was isolated using the Direct-zol™ RNA MiniPrep Kit (Zymo Research) according to manufacturer’s instructions, and the concentration determined by NanoDrop (Thermofischer). cDNA synthesis was performed using the iScript Advanced cDNAsynthesis kit (BioRad) according to manufacturer’s instructions. Subsequently, RT-qPCR was performed with a PCR mix containing 5 ng cDNA, 2.5 μl of SsoAdvanced SYBR qPCR mix (BioRad) and 0.25 μl of forward and reverse primers (250 μM concentration, Integrated DNA Technologies). The RT-qPCR was performed on a LC-480 device (Roche) and gene expression levels were analyzed using the qBase+ software 3.2 (Biogazelle). The software qBase+ uses a generalized model of the delta-delta-Ct approach, thereby supporting the use of gene-specific amplification efficiencies and normalization with multiple reference genes, with up to 204 genes. All formulas of this model are detailed in Hellemans et al., 2007. The RT-qPCR primers used in this study can be found in Table S12. Three biological and two technical replicates were collected per timepoint.

### Cell harvesting & protein extraction

The cells were scraped in PBS and collected in 15ml falcons. The collected cells were centrifuged at 4°C during 5 minutes at 2000rpm. The RIPA buffer (250 mg sodium deoxycholate, 40.5 ml water, 5M NaCl, 1M TrisHCl, 10% SDS solution, 10% NP-40) is used to lyse the cells for 45 minutes, following centrifugation (10min, 4°C, 8000g) the proteins were extracted. The concentration was determined using the BCA protein assay kit (Thermo Fisher, catalog number 23225).

### Immunoprecipitation followed by mass-spectrometry (IP-MS)

Protein from 15 to 20 million cells were extracted as mentioned above (Materials). Next, 2μg antibody (Anti-ZFHX4 antibody HPA023837, Rabbit (DA1E) mAb IgG XP® Isotype Control #3900S for WTIP vs IGGIP sample groups, respectively) was added and after four hours, 20 μl extensively washed protein A Ultralink resin beads (Thermo Scientific, catalog number 53139) were added, further rotated overnight. The samples were centrifuged and washed five times with 50mM digestion buffer (50 mM TrisHCl pH 8, 2 mM CaCl_2_) to remove unbound proteins. Beads containing antibody and proteins of interest were resuspended in 175 μl digestion buffer. For IP, 25 μl of samples were incubated at 95°C for 10 min with 6.25 μl of 5x laemmli buffer at 1000 rpm to elute the proteins. Supernatants containing the proteins of interest were used for western blot analysis. For IP-MS, the remaining 150 μl of the samples was incubated for 4 hours with 1 μg trypsin (Promega) at 37 °;C, while shaking. Beads were removed by centrifugation at maximum speed and proteins were digested again with 1 μg of trypsin, overnight at 37 °;C.

Peptides were re-dissolved in 20 µl loading solvent A (0.1% trifluoroacetic acid in water/acetonitrile (ACN) (98:0.5, v/v)) of which 2 µl was injected for LC-MS/MS analysis on an Ultimate 3000 ProFlow nanoLC system in-line connected to an Orbitrap Fusion Lumos mass spectrometer (ThermoFischer scientific). Trapping was performed at 20 μl/min for 2 min in loading solvent A on a 5 mm trapping column (ThermoFischer scientific, 300 μm internal diameter (I.D.), 5 μm beads). The peptides were separated on a 250 mm Aurora Ultimate, 1.7µm C18, 75 µm inner diameter (Ionopticks) kept at a constant temperature of 45°C in a butterfly heater (Phoenix S&T). Peptides were eluted by a non-linear gradient starting at 1% MS solvent B reaching 26.4% MS solvent B (0.1% FA in acetonitrile) in 30 min, 44% MS solvent B in 38 min, 56% MS solvent B in 40 minutes followed by a 5-minute wash at 56% MS solvent B and re-equilibration with MS solvent A (0.1% FA in water), all at a flow rate of 250 nl/min. The mass spectrometer was operated in data-independent mode, automatically switching between MS and MS/MS acquisition. Full-scan MS spectra ranging from 390-910 m/z with a target value of 4E5, a maximum fill time of 50 ms and a resolution at of 60,000 were followed by 30 quadrupole isolations with a precursor isolation width of 10 m/z for HCD fragmentation at an NCE of 34% after filling the trap at a target value of 4E5 for maximum injection time of 54 ms. MS2 spectra were acquired at a resolution of 30,000 at 200 m/z in the Orbitrap analyser without multiplexing. The isolation intervals were set from 400 – 900 m/z with a width of 10 m/z using window placement optimization.

QCloud^32,33^ has been used to control instrument longitudinal performance during the project.

### MS data analysis

LC-MS/MS runs of all samples were searched together using the DiaNN algorithm (version 1.9), library free. Spectra were searched against the human protein sequences in the Swiss-Prot database (database release version of 2022_01), containing 20,588 sequences (www.uniprot.org) and the imbedded protein contaminant database. Enzyme specificity was set as C-terminal to arginine and lysine, also allowing cleavage at proline bonds with a maximum of two missed cleavages. Variable modifications were set to oxidation of methionine residues and acetylation of protein N-termini. Mainly default settings were used, except for the addition of a 400-900 m/z precursor mass range filter, the match between runs (MBR) option. Further data analysis of the shotgun results was performed with an in-house script in the R programming language, version 4.2.2. Protein expression matrices were prepared as follows: the DIA-NN main report output table was filtered at a precursor and protein library q-value cut-off of 1 % and only proteins identified by at least one proteotypic peptide were retained. After pivoting into a wide format, iBAQ intensity columns were then added to the matrix using the DIAgui’s R package get_IBAQ function (https://rdrr.io/github/mgerault/DIAgui/man/DIAgui.html). PG.MaxLFQ intensities were log2 transformed and replicate samples were grouped. Proteins with less than three valid values in at least one group were removed and missing values were imputed from a normal distribution centered around the detection limit (package DEP)^34^ leading to a list of 2,795 quantified proteins in the experiment, used for further data analysis. To compare protein abundance between pairs of sample groups (WTIP vs IGGIP sample groups), statistical testing for differences between two group means was performed, using the package limma^35^. Statistical significance for differential regulation was set to a false discovery rate (FDR) of <0.05 and fold change of > 4- or < 0.25-fold (|log2FC| = 2). Results are presented in Supplementary Tables S8 and S9. Z-scored PG.MaxLFQ intensities from significantly regulated proteins were plotted in a heatmap after non-supervised hierarchical clustering. We analysed the significantly enriched genes from the MS data analysis with thousands of annotated gene set libraries retrieved from high-quality projects with the EnrichR tool^36–38^, and the results are visualized with a horizontal bar graph where the bars plotted horizontally show the gene sets found in different libraries and their corresponding pathways along the x-axis with the combined score with the measured p-values, q-values and z-scores which represents the measure of significance of enrichment level of the associated pathways.

### Western Blot

Western blot was performed according to an in-house protocol. Either protein lysates or eluted proteins after IP were subjected to NuPage gel (ThermoFischer Scientific, catalog number: EA0375BOX) electrophoresis, subsequently transferred to a nitrocellulose membrane, and detected using the appropriate antibodies: anti-ZFHX4 antibody (Sigma-Aldrich, HPA023837), anti-ZFHX4 antibody (Abnova, H00079776-M02), anti-vinculin (Merck Life Sciences, V9131). Blots were stripped twice. Imaging was performed with an Amersham Imager 600 CCD camera (GE Healthcare, Chicago, Illinois, USA).

### Zebrafish experiments

#### Breeding

Zebrafish lines were housed at the zebrafish facility Ghent in semi-closed recirculating housing systems (ZebTEC and WTU systems, Tecniplast) at a constant temperature (27-28°C), pH (∼7,5), conductivity (∼550µS) and light dark cycle (14/10). Zebrafish were fed every day with dry food (Gemma Micro) and with artemia (Ocean Nutrition). Zebrafish lines were maintained according to standard protocols^39,40^. Embryos/larvae were raised in E3 medium^41^ in a petri dish and placed in an incubator at 28°C, until 5 days-post-fertilization (dpf), the age at which experiments were performed.

#### zfhx4 crispant RNP complex generation & injections

We followed the protocol from Kroll et al, 2021^42^ for the generation of F0 knockout larvae for a single gene. Following the proper guidelines and by using CRISPOR^43^ we were able to find the best corresponding gRNAs for exons 2, 3 and 4 of *zfhx4*. The crRNAs were designed and produced by IDT (Integrated DNA Technologies) and checked in InDelphi^44^. The possible off targets with the highest probability found by CRISPOR, were assessed via PCR and sequencing on a Miseq instrument (Ilumina). Sequences of the *zfhx4* crRNAs, scrambled crRNAs and the corresponding primers used for genotyping of the injected embryos, are mentioned in Table S13.

To assemble the RNP, 2 nmol crRNA and 5 nmol tracrRNA pellets were respectively resuspended in 10 or 25 μL of the corresponding Duplex buffer (IDT), to obtain a 200 μM stock of each. These were stored at -20°C and were aliquoted to protect the RNA from the continuous thaw-freeze cycles and longer storage time. The crRNA was annealed with the tracrRNA to form the gRNA by mixing 1 μL crRNA with 1 μL of tracrRNA and 1,51 μL Duplex buffer. To obtain the duplex crRNA:tracrRNA (61 μM), this mix was heated to 95°C for 5 min, then cooled on ice. Each crRNA:tracrRNA complex was individually mixed with an equal molar amount Cas9. Each RNP complex was incubated 5 min at 37°C and had a concentration of 30.5 μM. The three RNP solutions were pooled in equal amounts (10.1 μM, 2 μL) making a total mix of 6 μL. The RNPs were stored at −20°C before the injections.

Around 1 nL for (first experiment) and 1 nL (second repeat experiment) of the three-RNP pool was injected into the single-cell yolk before cell inflation. This amounted to around 5029/800 pg of Cas9 and 1070/1000 pg of total gRNA. Each unique RNP was present in equal amounts in the pool. Therefore, in the case of three RNPs, 10.1/9.5 fmol of each RNP were injected in total with the Femtojet injector. The solution contained a dye to follow the successful injections.

#### Behavioral studies

Locomotor activity in response to light and dark stimuli was analyzed in zebrafish larvae (5dpf) using the DanioVision Observation Chamber (Noldus, The Netherlands) equipped with a Basler GenICam camera capturing 30 frames/second for image acquisition. Data was recorded and analyzed using the EthoVision XT 15 software (Noldus, The Netherlands). At 5 dpf, *zfhx4* crispants and scrambled zebrafish were transferred individually to a 24-well plate containing 1ml of E3 medium. Plates were subsequently placed in the DanioVision Observation Chamber with a protocol consisting of 20 min of 5% light (corresponding to 139 lux), 20 min dark, followed by alternating periods of 5 min 5% light and 5 min dark, for a total of 4 cycles. During the experiment, the temperature was kept constant at 28 °C using a Temperature Control Unit (Noldus). The first 10 minutes of the experiment were considered an acclimatization period and were not used for analyses. For each zebrafish, center-point detection was used. Movement frequency is depicted in time intervals of 1 min. This behavior experiment was repeated twice with n=60/genotype.

#### Genotyping

From each experimental and control condition, 30 5 dpf larvae were collected and DNA was extracted. The PCR reactions were performed on the exons of the sequence of interest (Table S13), following paired-end 2×150 bp sequencing on a Miseq instrument (Illumina) to determine the knockout efficiency following the protocol by Clement et al.^45^ The primers were used in the following PCR reaction: 2’ 94°C, 12× (30” 94°C, 30” 60*°C, 1’ 72°C), 25x (40” 94°C, 40” 48°C, 30” 72°C), 10’ 72°C) PCR reaction. The symbol “*” indicate in each cycle, lowering of the temperature by 1°C. The primers, target sequences and knockout efficiencies can be found in Table S13.

#### Alcian blue stainings and imaging

The following protocol is based on the published work from Stephan C.F NeuHauss^46^ and Delbaere et al., 2020^47^. Image analysis was performed to confirm the phenotype-genotype of crispants in comparison with scrambled zebrafish based on measuring different structures on the head of the 5 dpf injected larvae with the Leica fluorescent microscope M165 FC and a microscope light source. The magnification of the images was 12×. These stainings and measurements were repeated twice. We evaluated the length of the Meckel’s, the ethmoid plate, the distance between the middle point of the Meckel’s cartilage to the frontal point on the basihyal cartilage and the angle, the length of each ceratohyal cartilage and the angle between them with Fiji (Image J). Each measurement was normalized to the length of the middle point of both Meckel’s structures to the parachordal structure.

### DNA methylation analysis

DNA was obtained from the peripheral blood of individuals with *ZFHX4*-related syndrome and control subjects. Subsequently, a bisulfite-based technique was employed, and the levels of methylation were evaluated utilizing Illumina Infinium EPIC bead chip arrays (San Diego, CA, USA) following the prescribed procedures by the manufacturer. The assessment of sample quality, as well as the importation of normalized methylation and non-methylation signal intensity data, was conducted using the R minfi package (version 1.44.0)^48^.

The methylation data underwent analysis via a standard pipeline, outlined by Aref-Eshghi et al^49–51^, within the R environment (version 4.2.2). To ensure that disparities noted between the case and control cohorts were solely due to alterations in methylation rather than other potentially influencing factors, probes located on the X and Y chromosomes, probes prone to cross-reactivity with other genomic regions, and those with a detection p-value > 0.01 were removed from the analysis.

Probes exhibiting beta values of 0 and the top 1% most variable probes in terms of variance within either the case or control samples were excluded. Methylation levels, represented as beta values, underwent conversion into M-values through logit transformation. These transformed values were then employed for linear regression modeling utilizing the limma package (version 3.54.2)^35^.

The control samples were chosen from the EpiSign™ Knowledge Database (EKD)^49^ via the R software package MatchIt (version 4.5.2)^52^. Controls were matched by age, sex, and array type, resulting in the identification of 56 samples.

The model matrix was refined by incorporating estimated proportions of diverse blood cell types as covariates to address potential confounding influences^53^. Following this adjustment, p-values were refined for statistical significance using the eBayes function within the limma package.

The parameters for probe selection underwent adjustment to enhance the discrimination between the case and control samples. This evaluation utilized hierarchical clustering, employing Ward’s method based on Euclidean distance, and multidimensional scaling (MDS), which entailed scaling the pairwise Euclidean distances between samples for visualization purposes.

We employed the R package e1071 (version 1.7-13) to train a support vector machine (SVM) and construct a binary prediction model, following the methodology outlined in the referenced article^54^. The training process involved case samples, matched control samples, 75% of the remaining control samples and other rare genetic disorders from the EKD, while the remaining 25% of samples were reserved for testing purposes.

For each sample, the Methylation Variant Pathogenicity (MVP) score, ranging from 0 to 1, was computed to designate the similarity between its methylation pattern and the identified methylation profile for ZFHX4 case samples. The conversion of SVM decision values into MVP scores was accomplished using the Platt scaling method.

### CUT&RUN sequencing and data analysis

CUT&RUN was performed using Cutana pA/G-MNase (Epicypher, 15-1016) according to the manufacturer’s protocol. Briefly, cells (0.5M cells/sample) were washed, NE buffer was added to extract the nuclear protein and incubated with activated Concanavalin A beads for 10 min at room temperature. Cells were then resuspended in antibody buffer containing EDTA 0.5M, 1:100 dilution of each antibody (anti-ZFHX4 antibody, HPA023837; Rabbit (DA1E) mAb IgG XP® Isotype Control #3900S; anti-CTCF, Merck 07-729; H3K27ac antibody) was added to individual cell aliquots and tubes were rotated at 4 °C overnight. The following day, targeted chromatin digestion and release was performed with 2.5 mL Cutana pA/G-MNase and 100mM CaCl_2_. The genomic DNA was retrieved with the CUTANA™ DNA Purification Kit. Sequencing libraries were prepared with the NEBNext End Prep, following with the adaptor ligation and after size selection with Ampure Beads, the indexes were added, and the fragments are amplified. After the PCR amplification, the clean-up with the Ampure beads predominantly elutes mostly the fragments between 150-500 bp. The sequencing libraries were then sequenced with Illumina Nextseq 2000 using 2×150 bp paired-end reads to obtain 10 million read pairs per sample.

Read processing and peak calling was performed using a dedicated nextflow CUT&RUN pipeline available through nf-core^55^ (https://doi.org/10.5281/zenodo.5653535). Briefly, raw reads were trimmed, QC filtered and aligned to the hg38 reference genome using Bowtie2 (v2.4.4)^56^. Upon Q-score filtering, duplicate removal and CPM normalization, peaks in target samples were called with SEACR (v1.3)^57^ using corresponding IgG samples as controls. A consensus peak list per sample was obtained by requiring the presence of peaks in both replicates. Downstream annotation of consensus peaks was performed using the ChIPseeker R package^58^.

The top 20 of enriched GO terms (q value < 0.05) is visualized as a dot plot in Figure 6B. The Kyoto Encyclopedia of Genes and Genomes (KEGG) analysis provides the annotation of biological pathways, which is represented in Figure 6C. The top 10 of enriched pathways (q value < 0.05) is visualized as a dot plot. Homer37 (v4.11) was used to perform motif enrichment analysis, with 200 bp around the peak summit as input. The top 20 de novo and known motifs which were found from the analyses are summed up in Table S10.

### Human expression datasets

We examined the expression of ZFHX4 and its different isoforms in 54 non-diseased adult tissues (see Figure S1). To this end, the ZFHX4 expression levels in terms of TPM (transcripts per million) were retrieved from GTEX (https://www.gtexportal.org/home/). Furthermore, the ZFHX4 expression levels in terms of RPKM (reads per kilobase million) during neurodevelopment and adulthood were retrieved from Brainspan (http://www.brainspan.org, Figure 4). The brain-tissue-specific bulk RNA-seq data was downloaded from their website (“RNA-Seq Gencode v10 summarized to exons”). The brain regions were matched between development stage 2A and the later ones based on Table 2 in their technical white paper^59^. Read counts of the human embryonic craniofacial tissue bulk RNA-seq by Yankee *et al.* were accessed through their website (http://cotneyweb.cam.uchc.edu/craniofacial_bulkrna/)^60^. The ZFHX4 expression levels in human embryonic craniofacial tissue^60^ are represented in terms of RPKM (reads per kilobase million) from neurocranial crest cells to craniofacial tissue in embryonic periods.

The scRNA-seq neural organoid datasets^61^ (23 to 180 day of age) were accessed as RDS objects accessed through PsychENCODE on synapse (syn36139275), harmonized and integrated on a high performance computing unit (R version 4.1.0) using the “harmony” package (version 1.2.0)^62^. Plots were created using the ggplot2 package (version 3.5.2) in R (version 4.3.2)^63^. Lines in the plots were fitted using the ggplot2 function geom_smooth (method = “loess”) on the normalized counts (Figure 4C) or RPKM (Figure 4C). Plot from Figure 4D was created using the ggplot2 function “geom_bar()” with error bars made with stat_summary (fun.data = “mean_sdl”).

## Results

### Individuals harboring a *ZFHX4* deletion or protein truncating variant present with a clinically distinctive NDD

#### Demographic features

Data on 57 individuals (52 probands and 5 affected parents) with an aberration affecting *ZFHX4* (i.e. inversion, copy number variants (CNVs) and single nucleotide variants (SNVs) are summarized in Table S3 and more comprehensively documented in Table S1 and S2. There are 33 males and 24 females (Table S4). Ages ranged between five months and around their 50s. The median age at the last examination was around 8 years old.

#### Genotype

Twenty probands present with a structural variant affecting *ZFHX4*, including one chromosomal inversion and 19 CNVs. The inversion between genomic positions chr8:69893659 and chr8:76806725 (hg38) in proband 1 results in a breakpoint at chromosome 8q21.11 disrupting the *ZFHX4* intron between exons 4 and 5 (Figure 1). The CNVs are between 1.80 kb and 13.03 Mb in size, and comprise 13 multigenic deletions, six intragenic *ZFHX4* deletions (less than 90 kb in size), and one intragenic duplication that potentially causes a frameshift. The smallest region of overlap between the multigenic deletions is 361.9 kb (chr8:76444811-76806725 (hg38)) containing *ZFHX4* and *PEX2* (Figure 1A). Detailed information can be found in Table S1.

**Figure 1:**
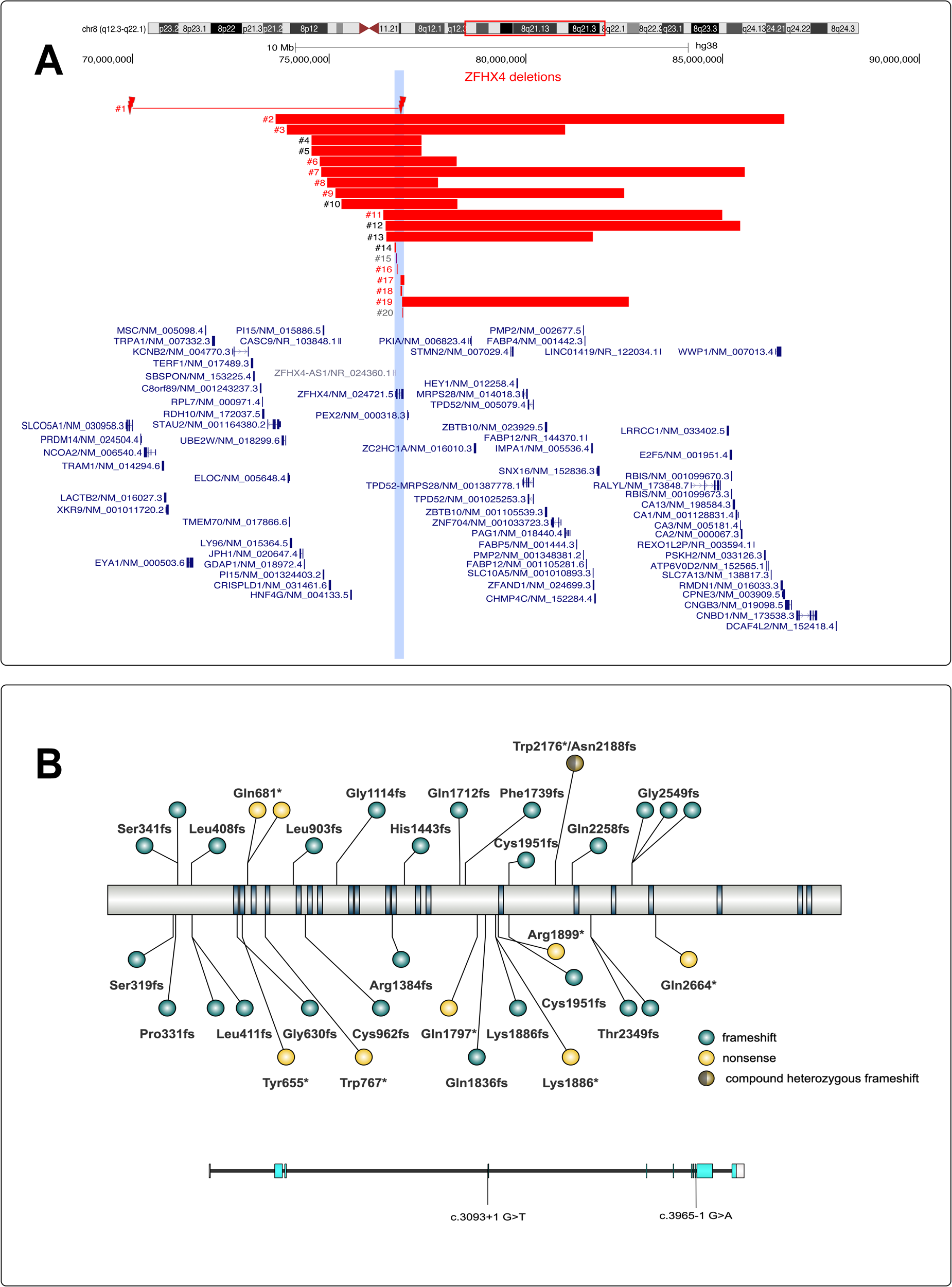
Overview of *ZFHX4* aberrations in 52 probands and 5 affected family members. (A) (Micro)deletions identified in probands 1–20 (#) are represented by the red bars and the duplication in proband 16 by the purple bar, all affecting *ZFHX4* (light blue box). RefSeq coding genes are indicated respectively in blue^131^. The numbers colored in black corresponding to their CNV, represent an individual with a training DNA sample for methylome analysis. The numbers colored in grey corresponding to each CNV variant, are individuals with their DNA samples used as testing samples for the methylome analysis. Genomic positions are according to hg38; *ZFHX4* is located on the reverse strand. (B) Nonsense, frameshift and splice variants in *ZFHX4* are identified in probands 21-52 and visualized with IBS 2.0^132^. Protein and gene structure of the canonical transcript (GenBank: NM_024721.5) are shown in the top and bottom part, respectively. The majority of PTVs are located in exon 2 and 10. Top: LoF variants are present in the ZFHX4 cohort and located in different domains of ZFHX4. Blue dots represent the frameshift variants, the yellow dots the nonsense variant and the brown-grey dot represents the variants of proband 52 which contains a nonsense (Trp2176*) and a frameshift (Asn2188*fs) mutation. The dot has been placed on the position of the nonsense variant (Trp2176*). Bottom: coding exons are indicated as light blue boxes and the UTRs are represented with white boxes.

Heterozygous frameshift, nonsense, and canonical splice site variants were identified in 23, seven, and two probands, respectively (Table S2). These PTVs were found in exon 2 (probands 21-30), exon 3 (probands 31 and 32), exon 5 (proband 34) and in the penultimate exon 10 (probands 36-51) and are all predicted to result in nonsense-mediated decay (i.e. > 55 bp away from last exon-exon boundary) (Figure 1B). All nonsense variants for which a CADD (combined annotation dependent depletion) score was available, have a score of 35 or higher, indicating that they are within the 0.1% most deleterious possible substitutions in the human genome. Probands 33 and 35 harbor a *ZFHX4* splice donor variant (*ZFHX4* (NM_024721.5):c.3093+1G>T) and a splice acceptor variant (*ZFHX4* (NM_024721.5):c3965-1G>A), respectively. According to SpliceAI^64,65^, the c.3093+1G>T variant results in loss of a canonical donor sequence (Figure S2) possibly leading to skipping of exon 3, while the c.3965-1G>A variant results in loss of the canonical acceptor sequence at exon 10 and gain of a novel acceptor causing a frameshift in exon 10 (Figure S2).

Additionally, we identified one individual (proband 52) who is compound heterozygous for a nonsense (*ZFHX4* (NM_024721.5):c.6528G>A /p.(Trp2176*)) and a frameshift variant (*ZFHX4* (NM_024721.5):c.6564_6567del/p.(Asn2188Lysfs*12)). Because the phenotypic consequences of the variants in probands 1 and 52 might be different, we excluded both individuals from the clinical characteristics analysis.

Twelve probands (4, 5, 8, 10, 15, 23, 29, 45, 47, 48, 49, and 52) inherited the variant from a similarly affected parent. The inheritance patterns of the *ZFHX4* variant in probands 21 & 50, mother of proband 23, mother of proband 29, mother of proband 47 could not be determined. In the remaining 38 probands, the aberration occurred *de novo* (i.e. 15 *de novo* CNVs and 23 *de novo* SNVs) (Table S1 & S2).

Only three variants (identified in probands 23, 25, 26 (same variant as proband 25) and 44) were found in the public variant database gnomAD (release v4.1.0). Twenty missense variants present in gnomAD affect the same codon as the truncating variant in probands 31, 32, 34, 36 38-41, 44, 46-52 (Table S5). While *ZFHX4* LoF variants appear to be under high constraint (as indicated by a pLI-score of 1 and a LOEUF score of 0.24), missense variants are not (as indicated by a Z score of 1.38), thereby suggesting that these missense variants do not result in LoF. Nevertheless, the presence of pathogenic variants in gnomAD does indicate that phenotypic variability may be wide.

#### Facial features

Dysmorphic facial features were reported in all individuals except for proband 21 for whom we have no data on the facial characteristics and probands 1 and 52 who were excluded from the clinical analysis (n=54/55; 98%). Frequently recurring facial characteristics are a broad lower half of the face (69%), a prominent forehead (71%), biparietal narrowing (47%), laterally sparse eyebrows (68%), eyelid ptosis (64%), periorbital fullness (53%), low-set ears (66%), prominent ear helices (66%), a wide nasal bridge (67%), flared nostrils (54%), underdeveloped nasal alae (64%) and an abnormal upper lip vermillion (51%). Facial characteristics observed in less than half of the individuals were hypertelorism (30%), upslanted palpebral fissures (44%), short palpebral fissures (37%), epicanthal folds (41%), posteriorly rotated ears (34%), a narrow nasal tip (41%), enlarged nares (29%), a smooth philtrum (34%), a thin upper lip (42%) and micrognathia (41%). Less frequently observed characteristics were microphthalmia (6%), a short philtrum (23%), an everted upper lip (21%), downturned corners of the mouth (20%), and cleft lip and/or palate (24%) (Table S3). A computationally based facial analysis using DeepGestalt facial analysis on 20 individuals with a *ZFHX4* deletion and 18 individuals with a LoF SNV in *ZFHX4* versus age-, and sex-matched controls, indicates a highly specific and sensitive gestalt with an average area under the curve (AUC) of 0.988 and 0.986 for individuals with a CNV or SNV, respectively, (p<0.005). Comparison of the facial features of both affected groups (i.e. *ZFHX4* deletion vs *ZFHX4* LoF variants) showed no significant difference (AUC=0.656 p value=0.131, figure not included).

#### Clinical findings

Global developmental delay and/or ID was present in most individuals (81%, n=39/48). However, in 7/31 individuals (23%), intellectual functioning was reportedly normal. For those with a documented IQ score (n=31), most had mild ID (n=16), two had borderline ID, five had moderate ID and one had severe ID. No statistical differences were found in the levels of ID between individuals with a CNV affecting *ZFHX4* and those with *ZFHX4* PTVs.

In Table S6, we included other features that were observed in our patients. More than half of individuals (n=30/47) presented with muscular hypotonia. Two third had behavioural difficulties (n=32/48), with stereotypies (n=8) and autism (n=10) being the most frequently reported. Morphological abnormalities of the central nervous system were found in 42% of the individuals, mostly ventriculomegaly (n=5), aplasia/hypoplasia of the corpus callosum (n=3) and widened subarachnoid space (n=3). Ophthalmological abnormalities included strabismus (48%, n=22/46), refraction abnormalities (49%, n=19/39) either myopia (n=8) or hypermetropia (n=5), nystagmus (n=1) and amblyopia (n=2). Notably, 8 individuals had an abnormal anterior eye segment morphology, with astigmatism (n=4), sclerocornea (n=2), microcornea (n=2), anterior segment dysgenesis (n=2), corneal opacity (n=1), and/or persistent pupillary membrane (n=1). Musculoskeletal manifestations included digital abnormalities in 45% (n=22/49) with clinodactyly (n=7) and camptodactyly (n=6) being most frequently reported. Growth abnormalities were described in 45% (n=20/45) of the probands. Prenatally, three individuals presented with intrauterine growth restriction and two with fetal macrosomia. Five individuals showed a postnatal growth retardation and short stature was documented in 27% (n=14/51). Genital abnormalities were found in 28% (n=11/40), including cryptorchidism (n=6), hypogenitalism (n=4) and abnormality of the scrotum (n=3). Congenital heart defects were reported in 17% (n=7/42) of the probands, mainly pulmonic stenosis (n=1), ventricular septal defect (n=1), atrial septal defect (n=2), aortic arch hypoplasia (n=1) and right aortic arch (n=1). About one third of the children (n=15/46) experienced feeding difficulties in infancy. Recurrent infections were noted in 17% (n=7/41).

#### Genotype-phenotype correlation

We compared the frequencies of 27 facial features and 15 clinical features between individuals with CNVs (group 1, n=20) and individuals with PTV variants in *ZFHX4* (group 2, n=34). Underdeveloped alae nasi (85% in CNVs, 54,5% in PTVs), abnormality of refraction (82% % in CNVs, 31% in PTVs), behavioral problems (87.5 % in CNVs, 58% in PTVs) and morphological abnormalities of the central nervous system (87.5 % in CNVs, 29% in PTVs) were more frequent in group 1 (p=0.023, p=0.004, p= 0.040 and p= 0.003 respectively), while cleft lip and or palate and micrognathia was frequent in group 2 (p=0.013and p= 0.030 respectively). ID is generally more severe in individuals with a multigenic deletion.

Furthermore, cleft lip and/or palate and prominent ear helix are more frequently observed in males versus females (i.e. 34% vs 9% for cleft lip/palate and 83% vs 50% for ear helix), while the penetrance of other clinical features did not differ between both sexes (p<0.05, Table S4).

### A 6.9 Mb *de novo* inversion involving chromosome bands 8q13-8q21.13 results in changes in 3D chromatin structure and disruption of *ZFHX4*

Individual 1 presented with a compatible but more severe phenotype (severe intellectual disability, significant dysmorphic facial features, abnormal muscle tone and abnormal heart morphology). We delineated the underlying 6.9 Mb *de novo* inversion spanning chromosome bands 8q13-8q21.13 at basepair resolution via genome sequencing (GS). This revealed an intergenic breakpoint on the centromeric end of the inversion and a breakpoint within intron 4 of *ZFHX4* on the telomeric right end (Figure 2). We next employed Hi-C to evaluate topologically associating domain (TAD) shuffling, revealing the typical bow tie configuration when mapped onto the reference genome (Figure 2). On the centromeric end, the inversion breakpoint resides within a TAD boundary (chr8:69875002-69900000 (hg38)), thereby possibly affecting two small (sub)TADs respectively containing the *SLCO5A1* and *PRDM14* gene. On the telomeric end of the inversion, the affected TAD contains *ZFHX4* and *PEX2* (Figure 2).

**Figure 2:**
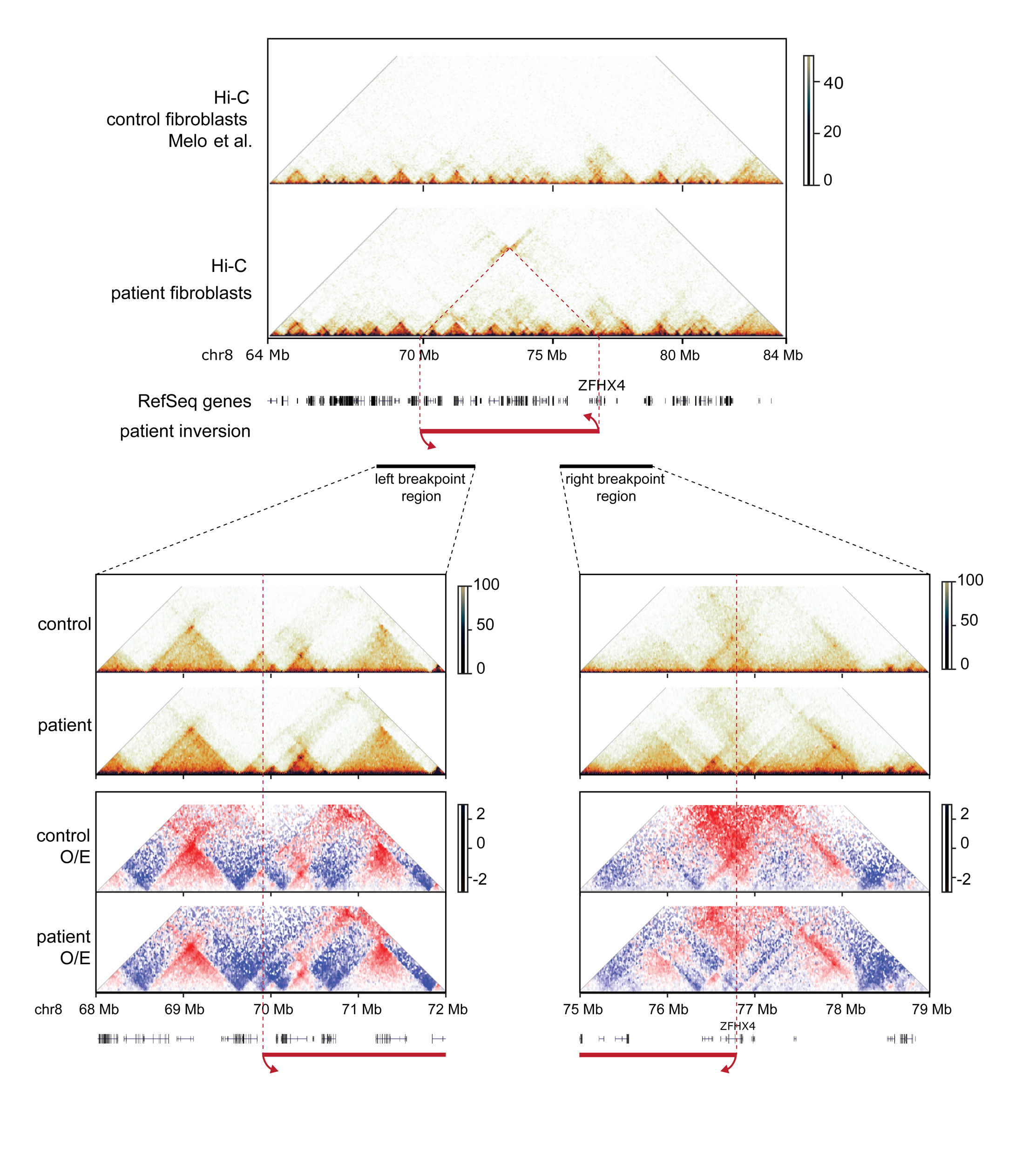
Hi-C on proband 1 fibroblasts reveals disrupted chromatin structure at inversion breakpoints. Hi-C contact frequency matrix for control and proband 1 fibroblast (inversion indicated by red arrows). Close-up of left and right breakpoint region, including observed/expected (O/E) contact frequencies for control and proband 1.

### Patients with a deletion encompassing a ‘critical region’ seem to show a mild methylation profile

Since we previously identified a methylation pattern associated with loss-of-function of *ZFHX3*^26^, a paralogue of *ZFHX4*, we also evaluated this for ZFHX4 loss-of-function. To this end, we performed methylome analysis on blood derived DNA samples of 26 individuals, including nine individuals with a CNV of *ZFHX4* (probands 4, 5 and their mothers, proband 10 and her father, probands 12-14), eleven individuals with a *ZFHX4* PTV variant (proband 23 and his mother, probands 24-25, 29, 30, 33, 35, 38, proband 45 and his mother), proband 15 with an intragenic *ZFHX4* duplication, proband 20 with an intragenic in-frame *ZFHX4* deletion and proband 1 with an inversion affecting *ZFHX4*, as well as three individuals from the cohort published in 2011^2^ (all listed in Table S7). No specific methylation difference was observed when comparing *ZFHX4* truncating variants with matched controls. However, DNA methylation analysis identified 227 CpG probes displaying notable methylation disparities between cases with deletions that included a minimal region (of 53 kb; deletion observed in proband 14) containing the *ZFHX4* promoter and matched controls, as evidenced by hierarchical clustering and MDS analyses (Figure 3A).

**Figure 3:**
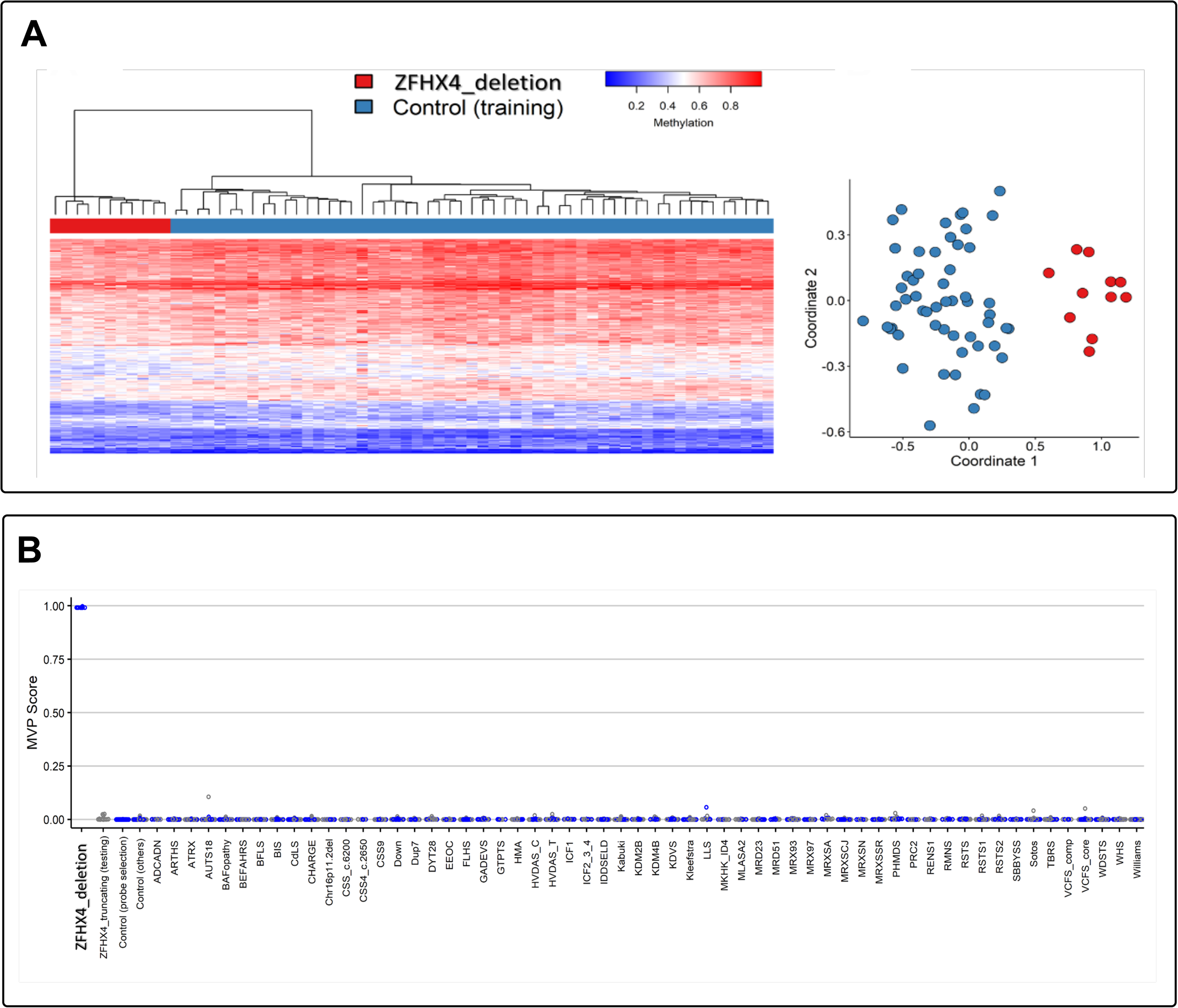
(A) The Euclidean clustering plot depicts ZFHX4 cases in red and the matched controls are shown in blue. The rows represent selected probes for the identified methylation pattern, while columns represent training cases and controls. A clear separation between cases and controls was observed. Methylation levels are color-coded: blue for 0 beta values and red for 1 beta values, with intensity indicating methylation level intensity. (**B) The MDS plot demonstrates the differentiation between case and control samples based on selected probes**. Cases are represented by red circles and controls by blue circles. C. Utilizing SVM classification trained with ZFHX4 samples against matched controls, as well as 75% of other control samples and samples from other disorders (blue), in addition to the remaining 25% of the database samples used for testing (gray), produced significant findings. Except for the *ZFHX4* deletion samples (training), all other conditions and ZFHX4 testing samples (*ZFHX4*_truncating) exhibited significantly low MVP scores, indicating an enhancement in the specificity of the model. The ZFHX4 testing samples, which encompass truncating variants (*ZFHX4*_truncating), a large duplication variant (*ZFHX4*_duplication), an inversion variant (*ZFHX4* _inversion) as well as an intragenic deletion (*ZFHX4*_in_frame_del) were included, enabling the model to predict class probability for ZFHX4 samples. Abbreviations: ADCADN, cerebellar ataxia, deafness, and narcolepsy, autosomal dominant; ARTHS, Arboleda-Tham syndrome; ATRX, alpha-thalassemia/intellectual development syndrome, X-linked; AUTS18, autism, susceptibility to 18; BEFAHRS, Beck-Fahrner syndrome; BFLS, Börjeson-Forssman-Lehmann syndrome; BIS, blepharophimosis intellectual disability SMARCA2 syndrome; CdLS, Cornelia de Lange syndrome1–4; CHARGE, CHARGE syndrome; 16p11.2del, 16p11.2 deletion syndrome; CSS_c.6200, Coffin-Siris syndrome (c.6232G>A [GenBank: NM_006015.4 (ARID1A)]; p.Glu2078Lys) (c.6254T>G [GenBank: NM_006015.4 (ARID1A)];p.Leu2085Arg) (c.6133T>C [GenBank: NM_017519.2 (ARID1B)]; p.Cys2045Arg); CSS4_c.2650, Coffin-Siris syndrome (c.2656A>G [GenBank: NM_001128849.1 (SMARCA4)]; p.Met886Val); CSS9, Coffin-Siris syndrome-9; Down, Down syndrome; Dup7, Williams-Beuren duplication syndrome (7q11.23 duplication syndrome); DYT28, Dystonia-28, childhood onset; EEOC, epileptic encephalopathy, childhood onset; FLHS, Floating-Harbour syndrome; GADEVS, Gabriele de Vries syndrome; GTPTS, Genitopatellar syndrome; HMA, HVDAS_C, Helsmoortel-Van der Aa syndrome (ADNP syndrome [central]); HVDAS_T, Helsmoortel-Van der Aa syndrome (ADNP syndrome [Terminal]); ICF1 Immunodeficiency, centromeric instability, facial anomalies syndrome 1, ICF2_3_4, immunodeficiency, centromeric instability, facial anomalies syndrome 2, 3, and 4; IDDSELD, intellectual developmental disorder with seizures and language delay; Kabuki, Kabuki syndromes 1 and 2; KDM2B, KDM2B-related syndrome; KDM4B, KDM4B-related syndrome; KDVS, Koolen de Vries syndrome; Kleefstra, Kleefstra syndrome 1; LLS, Luscan-Lumish syndrome; MKHK_ID4, Menke-Hennekam syndrome-1, 2; MLASA2, myopathy, lactic acidosis, and sideroblastic anemia-2; MRD23, intellectual developmental disorder, autosomal dominant 23; MRD51, intellectual developmental disorder, autosomal dominant 51; MRX93, intellectual developmental disorder, X-linked, XLID93; MRX97, intellectual developmental disorder, X-linked 97, XLID97; MRXSA, Armfield syndrome; MRXSCJ, syndromic X-linked intellectual disability, Claes-Jensen type; MRXSN, syndromic X-linked intellectual disability, Nascimento type; MRXSSR, syndromic X-linked intellectual disability, Snyder-Robinson type; PHMDS, Phelan-McDermid syndrome; PRC2, RENS1, Renpenning syndrome; RMNS, Rahman syndrome; RSTS, Rubinstein-Taybi syndrome-1, 2; RSTS1, Rubinstein-Taybi syndrome-1; RSTS2, Rubinstein-Taybi syndrome-2; SBBYSS, Say-Barber-Biesecker-Young-Simpson syndrome; Sotos, Sotos syndrome; TBRS, Tatton-Brown-Rahman syndrome; VCFS_comp, velocardiofacial syndrome; VCFS_core, velocardiofacial syndrome; WDSTS, Wiedemann-Steiner syndrome; WHS, Wolf-Hirschhorn syndrome; and Williams, Williams-Beuren deletion syndrome (7q11.23 deletion syndrome).

The support vector machine (SVM) classifier underwent training utilizing signature probes to precisely categorize individuals with ZFHX4-related syndrome, as observed in Figure 3B. It comprised 75% of control samples and other neurodevelopmental disorders (blue dots). The remaining 25% of control samples and other disorders were allocated for testing (grey dots). The *ZFHX4* samples with truncating variants (ZFHX4_truncating), the intragenic ZFHX4 duplication (ZFHX4_duplication), the inversion (ZFHX4 _inversion) and the in-frame deletion (ZFHX4_in_frame_del) exhibited low methylation variant pathogenicity (MVP) scores, which differed from the methylation pattern observed in the *ZFHX4* deletion samples (Figure 3B). Overall, we observed a distinct methylation profile for the deletion cases overlapping the deletion of proband 14, but not in individuals with intragenic *ZFHX4* defects nor LoF variants.

### *ZFHX4* expression increases during human neuronal differentiation, brain and craniofacial development

GTEx v8 (https://www.gtexportal.org/home/)) indicates abundant ubiquitous *ZFHX4* expression throughout the adult human body (Figure S1), except for several brain regions. However, expression profiles across seven organs (including brain and heart) through 23 developmental stages, show highest *ZFHX4* expression during the embryonic and fetal period, especially in the human brain and cerebellum^67^ (Figure S1). In line with these observations, we confirmed increased expression of *ZFHX4* in neural progenitor cells (NPCs) and early developed neurons upon *in vitro* neural differentiation starting from induced pluripotent stem cells (iPSCs) (Figure 4A). This increased expression was also confirmed at the protein level for NPCs in comparison to iPSCs (Figure 4B).

**Figure 4:**
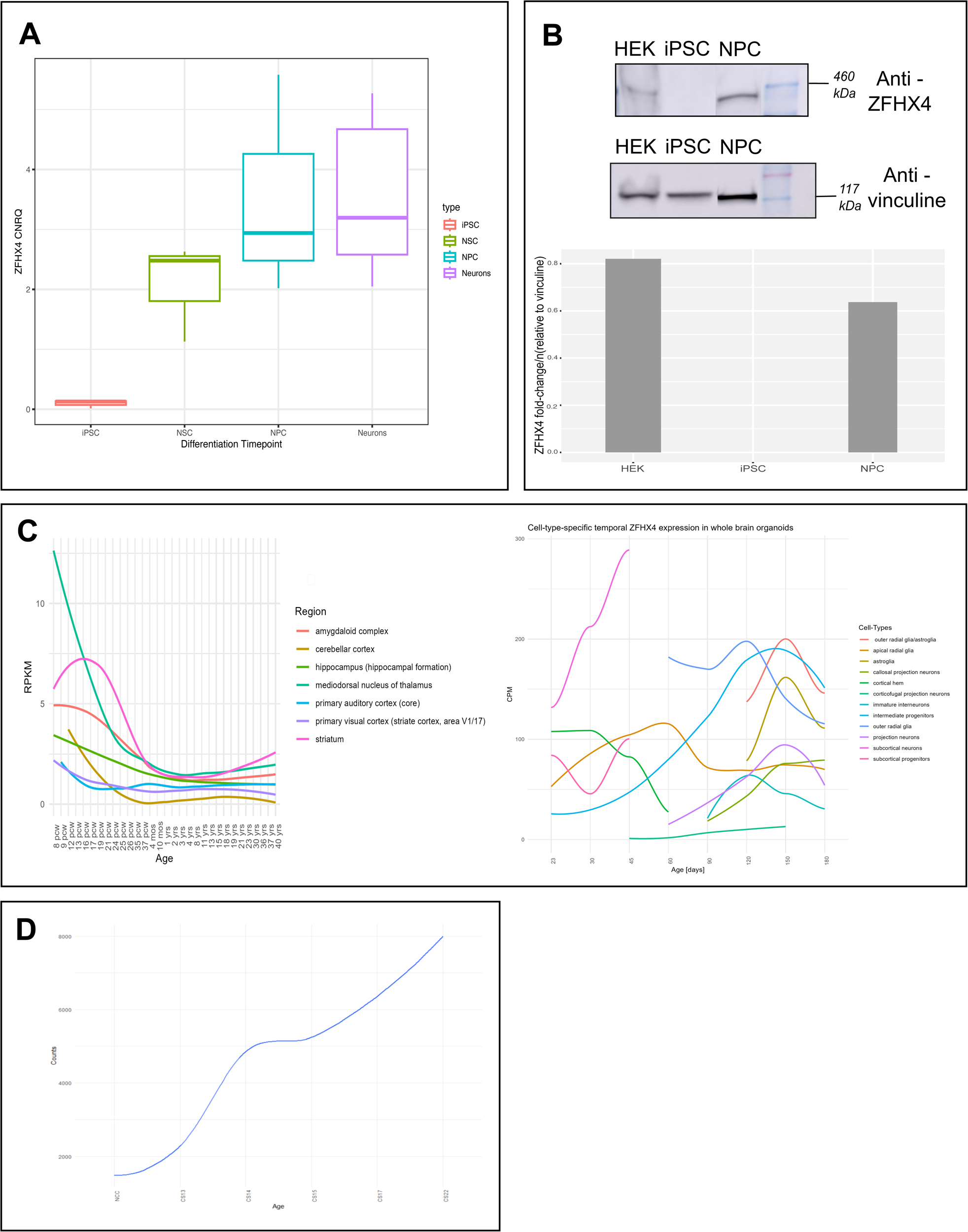
ZFHX4 expression increases during human neuronal differentiation, brain and craniofacial development (A) ZFHX4 expression levels during in vitro neural differentiation. RNA was sampled at the following stages: iPSCs, NSC (neural stem cells), neural progenitor cells (NPC) and direct differentiated neurons (Ns). An increasing expression is observed upon differentiation. The median expression value is indicated by the middle horizontal line. The whiskers indicate the minimum and maximum values. CNRQ, calibrated normalized relative quantity. **(B) Endogenous ZFHX4 (399 kDa) levels in HEK293T, iPSCs, and NPCs** as detected with western blot using the ZFHX4 antibody (top) and a vinculin antibody as a loading control (bottom). **(C) ZFHX4 levels in developing human brain.** ZFHX4 expression in the developing human brain (normalized RPKM data) showing higher expression during early prenatal development followed by decreased expression upon brain maturation. Data obtained from BrainSpan: http://www.brainspan.org. **(D) Human embryonic craniofacial tissue bulk RNA-seq** is visualized in a plot, showing an increase in ZFHX4 in craniofacial samples from five carnegie stages (CS13, CS14, CS15, CS17 and CS22)^133^.

Furthermore, reanalysis of transcriptomic data within the BrainSpan^59^ atlas of the developing human brain, also confirmed the highest *ZFHX4* expression in the brain between 8-16 weeks post conception, more specifically in the mediodorsal nucleus of the thalamus, striatum and the amygdaloid complex (left, Figure 4C). Through PsychENCODE, we retrieved single cell RNA-seq (scRNA-seq) datasets from developing neural organoids^61^. In the early differentiation stages between day 23 and 45, the highest *ZFHX4* expression is observed in the subcortical neurons (left, Figure 4C), while in day 120-150 neural organoids, this shifts to the outer radial glia and intermediate progenitors (right, Figure 4C). In addition, a clear increase in *ZFHX4* expression is observed in later embryonic stages of the developing facial bones^60^ (Figure 4D), in line with observed facial skeletal changes in patients with *ZFHX4* deficiency.

### ZFHX4 interacts with proteins involved in neural tube morphology and cellular organization in neural progenitor cells

Since NPCs showed increased ZFHX4 expression at the RNA and protein level, we performed immunoprecipitation followed by mass spectrometry (MS) in NPCs to discover potential interaction factors of ZFHX4. Forty-seven significantly enriched proteins were identified compared to an IgG isotype control (t-test FDR<0.05) (Figure 5A and B, Table S8 and S9). Biological process and pathway enrichment with EnrichR (https://maayanlab.cloud/enrichr-kg)^37,38^ revealed that ZFHX4 interacting proteins play a role in biological processes associated with histone modifications, development and cytosolic transport (Figure 5C). Furthermore, ZFHX4 interacts significantly with proteins encoded by genes associated with neurodevelopmental disorders: *ZMIZ1 (*MIM#: 618659), *ZEB1* (MIM#:613270 & 609141)*, WDR81* (MIM#: 610185 & 617967)*, KCNQ2* (MIM#: 613720 & 121200)*, KMT2D* (MIM#: 620186 & 147920)*, NUP107* (MIM#: 618348 & 616730), *IMPA2*^68^, *ZMYND8*^69^, *NACC1* (MIM#: 617393) and *WASHC4* (MIM#: 615817).

**Figure 5:**
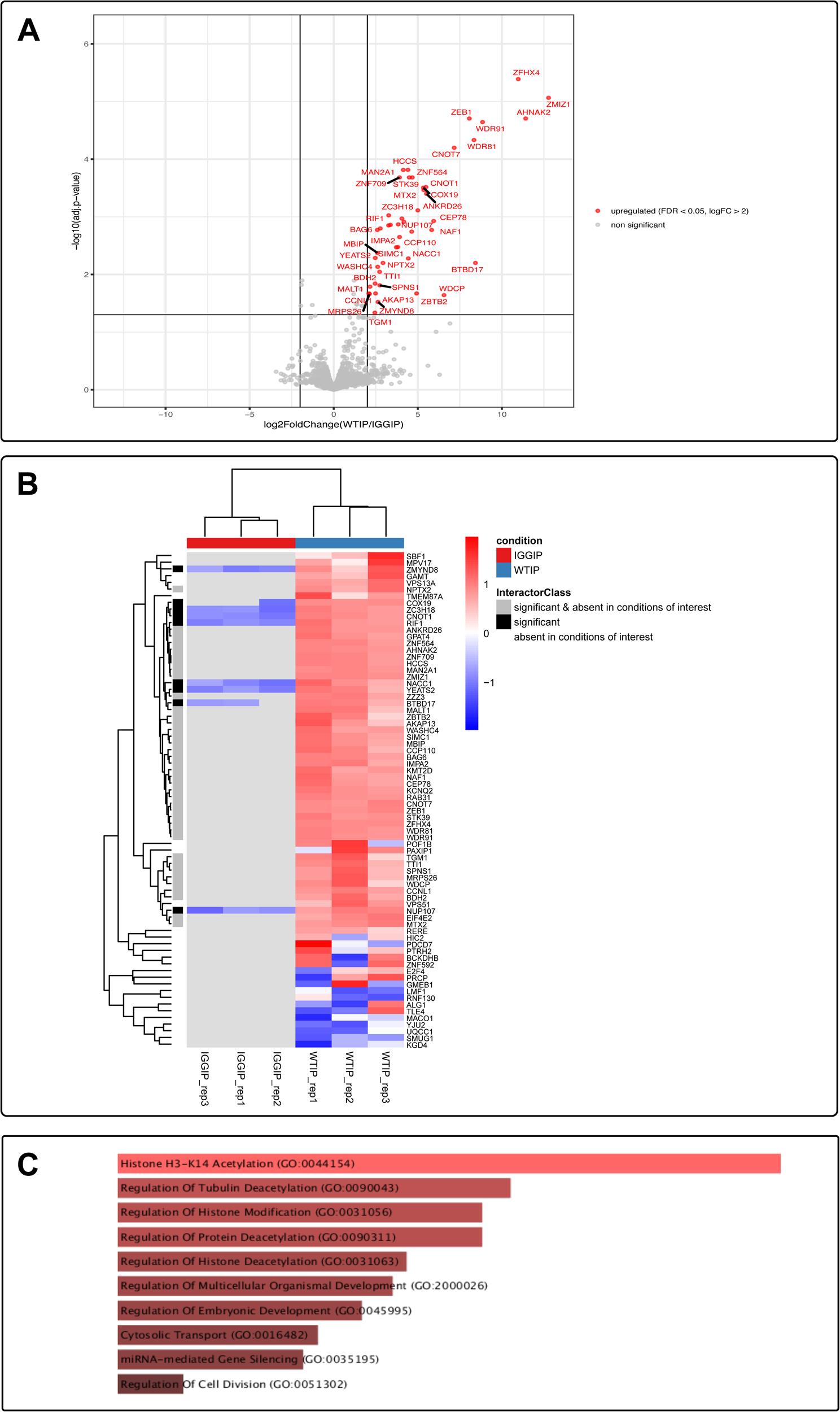
ZFHX4 interacts with 46 enriched proteins involved in axonogenesis, cell differentiation and neuron differentiation. (A) The enriched ZFHX4 interacting proteins in NPCs shown in a volcano plot. For each detected protein, the log2 fold change (FC) between the ZFHX4 IP-enriched samples and the IgG controls is shown on the x axis, while the -log10 of the p value is shown on the y axis. The horizontal dashed line indicates the -log10 value of a p value equal to 0.05. Proteins with an FDR < 0.05, log2 FC > 2 and log2 FC < -2 are, respectively, highlighted in blue and red. **(B) Hierarchical clustering of potentially interesting proteins.** The proteins shown to be differentially abundant between groups are visualized in a heatmap after non-supervised hierarchical clustering of z-scored log 2 PG.MaxLFQ intensities. The heatmap color scale from blue to red represents the level of abundance. The interacting factors which are absent in the IgG control, significant but not absent at the IgG control and significantly enriched protein and absent in the IgG control, are listed in Tables S7 and S8. **(C) ZFHX4 interacting factors associated to their biological processes with EnrichR.**

### ZFHX4 binds to the promoter of genes involved in axonogenesis, regulation of the nervous system development, neurogenesis and neural migration

To assess the downstream target genes of the transcription factor ZFHX4, we performed CUT&RUN for ZFHX4 on two NPC replicates, retrieving around 1640 consensus peaks mainly consisting of promoter regions (55.62%) (Figure 6A & Table S10). Furthermore, Gene Ontology analysis of these peaks showed that ZFHX4 binds to the promoter of genes involved in (the regulation of) axonogenesis, forebrain development, cell differentiation nervous system development and neuron differentiation (Figure 6B). KEGG pathway analysis indicated that the proteins encoded by these genes are involved in axon guidance, the Wnt, Rap1 and other signaling networks regulating pluripotency of stem cells. All pathways are strongly involved in cell organization, embryonic and nervous system development (Figure 6C)^70–76^. Motif enrichment analysis of the consensus peaks revealed an enrichment of homeobox, transcription factors, SOX-related genes and zinc finger motifs (Table S11).

**Figure 6:**
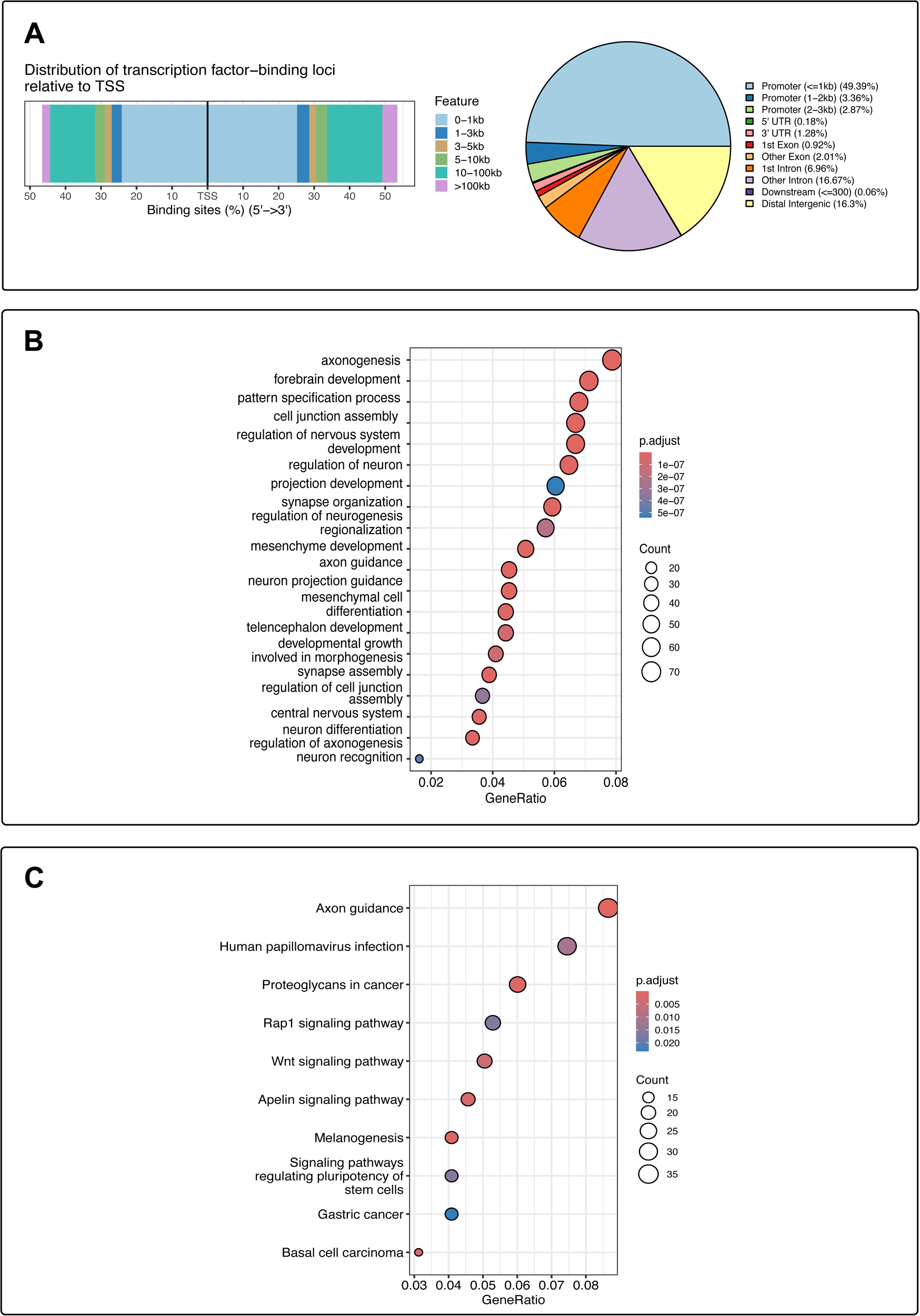
CUT&RUN sequencing data shows ZFHX4 binding to genes involved in axonogenesis, the regulation of the nervous system development, neurogenesis and neural migration. (A) Annotation of CUT&RUN consensus peaks reveals ZFHX4 binding to promoter regions. Left: Distribution of the ZFHX4 consensus peaks relative to transcription start sites (TSS). Half of the peaks are found between 0-1 kb of the TSS. Right: Pie chart visualizing the genomic annotation and the percentage of the peaks that overlap a TSS, 5’ UTR, 3’ UTR, exonic, intronic, downstream or distal intergenic regions. This shows that 55.62% of ZFHX4 consensus peaks are found near gene promoters. **(B) ZFHX4 binds to genes involved in axonogenesis, forebrain and nervous system development, including the regulation of the neurogenesis and synapse organization; cell and neuron differentiation.** The gene ontology (GO) dot plot displays the top 20 enriched biological processes (BPs) ranked by gene ratio (the number of genes related to GO term/total number of significant genes) and the p-adjusted values for these terms (colour, p-value 1e10^-7^-1e10^-5^). The size of the dot represents the gene counts per BP. **(C) ZFHX4 plays a key role in embryonic, neuron and axon developmental pathways.** The Kyoto Encyclopedia of Genes and Genomes (KEGG) dot plot provides all enriched biological pathways ranked by gene ratio (# of genes related to GO term/total number of significant genes) and the p-adjusted values for these terms (color). The size of the dot represents the gene counts per pathway.

### zfhx4 F0 zebrafish crispant larvae show craniofacial and behavioral phenotypes

The zebrafish *zfhx4* gene (ENSDARG00000075542) resides on chromosome 24, contains 10 exons and has 71.37% sequence identity with the human *ZFHX4* (Figure 7A). In 2021, Kroll *et al.* showed that first-generation (F0) mosaic crispant zebrafish recapitulate the phenotype of germline knock-outs^42^. We generated *zfhx4* crispants by injection of a mixture of three crRNAs obtaining a *zfhx4* indel efficiency of approximately 79% (see Table S13).

**Figure 7:**
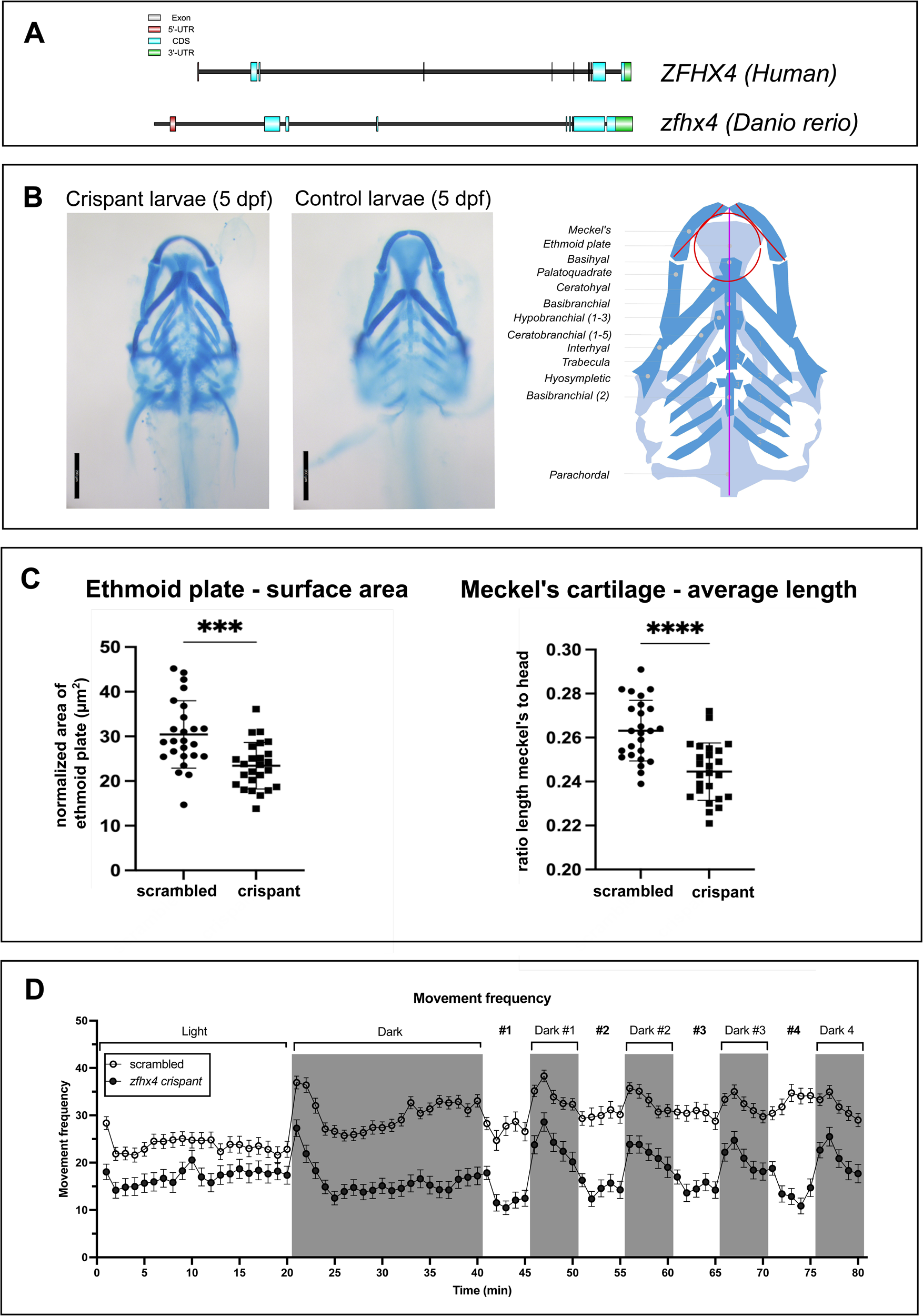
Zebrafish *zfhx4* crispants showed both craniofacial and behavioral changes. (A) Gene structure of the zebrafish *zfhx4* and *ZFHX4*. Top: The coding exons of *ZFHX4* are shown as light grey boxes and light blue boxes for the CDS (coding sequence), and untranslated regions in red (5’-UTR) and green (3’-UTR). Bottom: The coding exons of *zfhx4* are shown as light grey boxes and light blue boxes for the CDS, and untranslated regions in red (5’-UTR) and green (3’-UTR). **(B) Representative craniofacial images for *zfhx4* and scrambled zebrafish crispants are shown**. Left: microscopic ventral view scrambled (top panel) and *zfhx4* crispants (bottom panel) of Alcian blue stained craniofacial structures at 5 dpf reveal smaller ethmoid plates and shorter Meckel’s cartilage in the F0 *zfhx4* crispant larvae. Right: Adapted image^46^ from the representation of the cartilage structures of the zebrafish neurocranium (dark blue) at 5 dpf by lines. The red lines and circle show the measurements taken and normalized with the length of the head (purple line). **(C) Left: the surface area of the ethmoid plate was measured and normalized to the head length of the larvae. Right: The length of the individual Meckel’s cartilage structures was measured and normalized to the head length of the larvae.** The ethmoid plate and the Meckel’s cartilage showed significant differences between crispant larvae compared to scrambled larvae (p-value ≤ 0.001 (***), p-value ≤ 0.0001 (****)) **(D) *zfhx4* crispants show a consistently lower movement frequency compared to scrambled control larvae during the complete behavior experiment but the trends during light and dark cycles remain similar**. Dark periods are indicated with grey boxes. Data is represented as mean ± SEM (n=60/genotype).

To determine craniofacial cartilage defects, we stained neural crest-derived chondrocytes in 5dpf *zfhx4* crispant larvae and scrambled crRNA control larvae with Alcian Blue (Figure 7B). *zfhx4* F0 crispant larvae had a significantly smaller ethmoid plate and shorter Meckel’s cartilages in comparison with scrambled larvae ((p-value ≤ 0.001 (***), p-value ≤ 0.0001 (****), Figure 7C), thereby confirming that ZFHX4 LoF results in craniofacial anomalies.

Next, locomotor behavior was assessed in response to light/dark transitions using the Daniovision Observation Chamber. Analysis of the swimming pattern of scrambled zebrafish showed a higher movement frequency during the dark cycles. *Zfhx4* crispant zebrafish showed a movement pattern comparable to scrambled zebrafish during the light and dark cycles, but with a consistently lower movement frequency (Figure 8D and Figure S6, p-value ≤ 0.05 (*), p-value ≤ 0.0001 (****)).

## Discussion

In this study, we delineated a clinically distinctive neurodevelopmental phenotype caused by pathogenic variants in *ZFHX4*. Most individuals presented with mild to moderate intellectual disability and behavioral problems, often associated with anatomical brain anomalies. All assessed individuals showed a recurrent facial gestalt. Short stature, digital anomalies, genital defects, cleft lip/palate, ophthalmological anomalies and feeding difficulties were present in 25-50% of the affected individuals, while congenital heart defects were occasionally observed. The high phenotypic similarity, including the facial characteristics, between individuals with CNVs including *ZFHX4* and individuals with intragenic *ZFHX4* variants indicates that *ZFHX4* loss-of-function is the mechanism underlying the common phenotype. However, individuals with CNVs appear to have an increased risk for behavioral abnormalities (Fisher’s exact test p=0.0402), while the prevalence of cleft lip/palate was lower compared to individuals with PTV variants in *ZFHX4*. Furthermore, the methylation profile identified in individuals with 8q21.11 (micro)deletions encompassing *ZFHX4* could not be replicated in individuals with PTVs. Although other genes contained in the 8q21.11 (micro)deletions (Table S14) might contribute to or modify the methylation pattern, the smallest region of overlap among all deletion samples tested for a methylome signature, is the size of the deletion in individual 14 (53 kb). This deletion does not only partially delete *ZFHX4*, but also removes a non-coding region containing the *ZFHX4* promoter. In the other tested individuals with a *ZFHX4* PTV, an intragenic duplication, an intragenic deletion or an inversion, the aberration does not result in the removal of this critical region. This suggests that deletion of this critical region might be the main contributor to its pattern. To confirm a possible role for this ‘critical’ region as one of the drivers for the methylation profile, analysis of additional individuals is necessary.

Of note, twelve variants (i.e. six CNVs and six SNVs) were inherited from healthy parents, of whom three also had learning difficulties in childhood, although without a formal diagnosis. Although *ZFHX4* is very intolerant for LoF variants (pLI=1; LOEF=0.24), 172 LoF variants (excluding low coverage ones) are present in gnomAD (v4.1.0), including three variants present in our cohort (albeit all with a very low frequency in the general population). As we observed a wide phenotypic spectrum in individuals with *ZFHX4* CNVs and PTVs, analysis of *ZFHX4* should not be limited to the identification of *de novo* aberrations. Moreover, careful recording of the phenotypic data of the parents remains important.

Individual 1 harbors a *de novo* inversion disrupting *ZFHX4* and presented with a typical but more severe phenotype. Via Hi-C, we observed that the inversion affects two small (sub)TADs on the centromeric end, containing *SLCO5A1* and *PRMD14* gene. At the telomeric end, the affected TAD contains *ZFHX4* and *PEX2* (Figure 2). *SLCO5A1* and *PEX2* have been previously linked to disease. *PEX2* is associated with the peroxisome biogenesis disorder 5A and 5B, both autosomal recessive multiple congenital anomaly disorders (MIM 614866 & 614867). *SLCO5A1* on the other hand, has been linked to the autosomal dominant Mesomelia synostoses syndrome (MSS, MIM: 600383)^79,80^. MSS is characterized by mesomelic shortening of the limb, acral synostoses and multiple congenital malformations. In 2010, Isidor *et al.* reported 8q13 deletions associated with MSS^81^. Although these deletions varied in size, they always encompassed *SULF1* and *SLCO5A1*. Furthermore, Roshandel *et al.* generated an siRNA mediated knockdown of *Oatp30B* (i.e., the Drosophila orthologue of *SLCO5A1*) and observed a startling and seizure-like phenotype^82^. Although *SLCO5A1* is not directly affected in individual 1, the inversion might exert an effect on the expression of *SLCO5A1*, thereby combining both the ZFHX4 and MSS phenotypes. However, further functional studies are necessary to unravel this.

Although previous reports suggested *ZFHX4* as a candidate gene for non-syndromic orofacial clefting^10^, we observed orofacial clefting in only about 30% of the individuals with *ZFHX4* LoF variants. We investigated the craniofacial cartilage development in mosaic knockout *zfhx4* zebrafish and observed significant differences between the Meckel’s cartilage and the ethmoid plate structures that correspond to the human jaw and palate, respectively. A homozygous *Zfhx4* knock-out mice model further supports this hypothesis, presenting cleft palate at 100% penetrance^17^. In addition to the craniofacial differences in our zebrafish model, we also observed behavioral changes. The decreased movement frequency could be attributed to (a combination of) neurological, skeletal and/or muscle defects in the *zfhx4* crispant zebrafish larvae, given the role of ZFHX4 in different biological processes of these tissues^12,14,17^. Interestingly, the International Mouse Phenotyping Consortium (IMPC) applied a panel of phenotyping screens on *Zfhx4* homozygous and heterozygous knock-out mice. Similar to what was observed by Nakamura *et al.*, the homozygous *Zfhx4* deficient mice showed full penetrant preweaning lethality. However, the heterozygous *Zfhx4* deficient mice were viable (11 females and 13 males), but did not present with cleft palate, and showed decreased exploration in a new environment. Our *in vivo* zebrafish results as well as the previous studies in mice, clearly indicate a relationship between *ZFHX4,* clefting and behavioral abnormalities.

The complex structure of ZFHX4, containing 4 homeodomains and 22 zinc fingers, suggests dynamic functions in diverse biological processes. Our IP-MS screen identified ZFHX4 protein interactors involved in the regulation of histone modifications, development, transport, gene expression and cell division. Most of the interacting factors are associated with histone modifications and development. YEATS2, ZZZ3, KMT2D and ZMYND8 play a crucial role in chromatin remodeling, histone modifications, such as acetylation^86–92^. Although CHD4, a crucial member of NuRD complex, was previously identified as ZFHX4 interacting protein in glioblastoma cells, it was not identified as an interaction partner in our NPC screen. However, we identified the CHD4-interacting multivalent histone reader ZMYND8 as interacting factor of ZFHX4. Furthermore, in 2022, Dias *et al.* reported disruption of ZMYND8 as a cause for syndromic intellectual disability^69^. Moreover, several of the other ZFHX4 interacting proteins have also been associated with NDDs: YEATS2 plays an important role in histone acetylation and has been involved with epilepsy (MIM: 615127; epilepsy, myoclonic, familial adult)^87,88^. ZEB1 has been associated in corneal dystrophy (MIM: 613270 and 609141)^93,94^. Another interacting factor is *KCNQ2* (MIM: 613720; developmental and epileptic encephalopathy), encoding a potassium voltage-gated channel, which plays a key role in the regulation of neuronal excitability^95,96^. Mutations in *KCNQ2* have been associated with self-limited familial neonatal epilepsy (SelFNE) and other forms of epilepsy (early-onset developmental and epileptic encephalopathy)^97,98^. Additionally, mutations in *KMT2D*, encoding a histone methyltransferase give rise to Kabuki syndrome (MIM:147920, Kabuki syndrome 1)^99,100^, associated to neural crest cell differentiation and facial morphology^101,102^. IMPA2 plays an important role in phosphatidylinositol signaling pathway and has been associated with susceptibility to bipolar disorder^68,103^. Furthermore, GAMT is crucial for creatine biosynthesis and associated with cerebral creatine deficiency syndrome 2 (MIM:612736)^104–106^. Other interacting factors, such as NPTX2 and ZNF592, are involved in synaptic plasticity and neuronal development^107–110^.

Furthermore, in NPCs, ZFHX4 mainly binds to the promoter regions of genes that are significantly enriched for homeobox, SOX-related genes and zinc fingers motifs according to *de novo* and known HOMER motif analysis (Table S11). Via SEA motif enrichment, the same consensus peaks seem to show ERF-, KLF (Krüppel-like factors) and SP (specificity protein) motif enrichment (Table S11). These transcription factors are crucial for transcriptional regulation and cell proliferation^83,84^. Mutations in ERF genes have been associated with craniosynostosis with facial dysmorphism^62^, while KLF and SP genes are involved in embryonic development and tissue differentiation^83^.

In conclusion, we have delineated the ZFHX4-related NDD and identified ZFHX4 haploinsufficiency as a cause of autosomal dominant syndromic intellectual disability. Our initial findings indicate an important role for ZFHX4 in craniofacial and brain development, through interaction with proteins involved in chromatin organization.

## Web Resources

UniProt website: www.uniprot.org

Evo-devo mammalian organs app: https://apps.kaessmannlab.org/evodevoapp/

GTEx portal browser: https://www.gtexportal.org/home/

BrainSpan Atlas of the Developing Human Brain: http://www.brainspan.org

Human embryonic craniofacial tissue bulk RNA-seq website: http://cotneyweb.cam.uchc.edu/craniofacial_bulkrna/

Human Organoid Single-Cell Browser (Shcheglovitov Lab): https://shcheglovitov.shinyapps.io/u_brain_browser/gnomAD

GnomAD v4.1.0: https://gnomad.broadinstitute.org/about

GTEx v8: https://www.gtexportal.org/home/

## Data Availability

The data will be published in public servers after publication.

## Acknowledgements

We extend our gratitude to the participating families for their contribution in this study. We also appreciate the collaboration with the VIB Proteomics Core, led by Prof. Francis Impens. We are particularly grateful for the data analysis conducted by the bioinformatics team, specifically Simon Devos and Sara Dufour. Additionally, the zebrafish project was made possible with the support of the Zebrafish Facility Ghent Core Facility at the Center for Medical Genetics Ghent (Ghent University), with special thanks to Andy Willaert and Hanne De Saffel.

Financial support has been provided by grants 1520518N and G055422N from the Research Foundation – Flanders (FWO) and BOF/STA/201909/009 from the Special Research Fund (BOF) from Ghent University. M.d.R.P.B is supported by a doctoral grant (STI.DIV.2023.0005.01) of the Marguerite-Marie Delacroix foundation. B.C. is a senior clinical investigator of the Research Foundation Flanders.

Research on proband 20 was possible by access to the data generated by the France Genomic Medicine Plan 2025.

The work generated for proband 27 was performed within the European Reference Network for Intellectual disability, TeleHealth, Autism and Congenital Anomalies (ERN ITHACA).

Sequencing and analysis of probands 35 and 45 were provided by the Broad Institute of MIT and Harvard Center for Mendelian Genomics (Broad CMG) and were funded by the National Human Genome Research Institute (NHGRI) grants UM1HG008900 (with additional support from the National Eye Institute, and the National Heart, Lung and Blood Institute), U01HG0011755, R21HG012397, and R01HG009141, and in part by the Chan Zuckerberg Initiative Donor-Advised Fund at the Silicon Valley Community Foundation (funder DOI 10.13039/100014989) grants 2019-199278, 2020-224274, 2022-316726 (https://doi.org/10.37921/236582yuakxy).

Angela Peron would like to thank ASST Santi Paolo e Carlo, San Paolo Hospital for the support during this study.

## Data and code availability

The analyzed IP-MS data and the ZFHX4 consensus peaks of the CUT&RUN sequencing, as well as the results of the *de novo* and known motif analyses are available in the supplemental information.

## Supplementary material

Supplemental information can be found online.

## References

1. Jansen, S., Vissers, L.E.L.M., and de Vries, B.B.A. (2023). The Genetics of Intellectual Disability. Brain Sci 13, 231. 10.3390/brainsci13020231.

2. Palomares, M., Delicado, A., Mansilla, E., De Torres, M.L., Vallespín, E., Fernandez, L., Martinez-Glez, V., García-Miñaur, S., Nevado, J., Simarro, F.S., et al. (2011). Characterization of a 8q21.11 microdeletion syndrome associated with intellectual disability and a recognizable phenotype. American Journal of Human Genetics 89, 295–301. 10.1016/j.ajhg.2011.06.012.

3. Hofmann, K., Becker, J., Heller, R., Boute, O., Andrieux, J., Hoyer, J., Ekici, A.B., Reis, A., and Rauch, A. (2011). 7 Mb *de novo* deletion within 8q21 in a patient with distal arthrogryposis type 2B (DA2B). European Journal of Medical Genetics 54, e495–e500. 10.1016/j.ejmg.2011.06.002.

4. Vulto-van Silfhout, A.T., Hehir-Kwa, J.Y., van Bon, B.W.M., Schuurs-Hoeijmakers, J.H.M., Meader, S., Hellebrekers, C.J.M., Thoonen, I.J.M., de Brouwer, A.P.M., Brunner, H.G., Webber, C., et al. (2013). Clinical significance of de novo and inherited copy-number variation. Hum Mutat 34, 1679–1687. 10.1002/humu.22442.

5. Happ, H., Schilter, K.F., Weh, E., Reis, L.M., and Semina, E.V. (2016). 8q21.11 microdeletion in two patients with syndromic peters anomaly. Am J Med Genet A 170, 2471–2475. 10.1002/ajmg.a.37840.

6. Belligni, E.F., and Hennekam, R.C.M. (2010). Familial occurrence of ptosis, nasal speech, prominent ears, hand anomalies and learning problems. European Journal of Medical Genetics 53, 192–196. 10.1016/j.ejmg.2010.03.009.

7. Li, S., and Hao, G. (2021). Current Trends and Prospects in Compliant Continuum Robots: A Survey. Actuators 10, 145. 10.3390/act10070145.

8. Eising, E., Carrion-Castillo, A., Vino, A., Strand, E.A., Jakielski, K.J., Scerri, T.S., Hildebrand, M.S., Webster, R., Ma, A., Mazoyer, B., et al. (2019). A set of regulatory genes co-expressed in embryonic human brain is implicated in disrupted speech development. Mol Psychiatry 24, 1065–1078. 10.1038/s41380-018-0020-x.

9. Kaplanis, J., Samocha, K.E., Wiel, L., Zhang, Z., Arvai, K.J., Eberhardt, R.Y., Gallone, G., Lelieveld, S.H., Martin, H.C., McRae, J.F., et al. (2020). Evidence for 28 genetic disorders discovered by combining healthcare and research data. Nature 586, 757–762. 10.1038/s41586-020-2832-5.

10. Bishop, M.R., Diaz Perez, K.K., Sun, M., Ho, S., Chopra, P., Mukhopadhyay, N., Hetmanski, J.B., Taub, M.A., Moreno-Uribe, L.M., Valencia-Ramirez, L.C., et al. (2020). Genome-wide Enrichment of *De Novo* Coding Mutations in Orofacial Cleft Trios. The American Journal of Human Genetics 107, 124–136. 10.1016/j.ajhg.2020.05.018.

11. Karczewski, K.J., Francioli, L.C., Tiao, G., Cummings, B.B., Alföldi, J., Wang, Q., Collins, R.L., Laricchia, K.M., Ganna, A., Birnbaum, D.P., et al. (2020). The mutational constraint spectrum quantified from variation in 141,456 humans. Nature 581, 434–443. 10.1038/s41586-020-2308-7.

12. Hemmi, K., Ma, D., Miura, Y., Kawaguchi, M., Sasahara, M., Hashimoto-Tamaoki, T., Tamaoki, T., Sakata, N., and Tsuchiya, K. (2006). A Homeodomain-Zinc Finger Protein, ZFHX4, Is Expressed in Neuronal Differentiation Manner and Suppressed in Muscle Differentiation Manner. Biological & Pharmaceutical Bulletin 29, 1830–1835. 10.1248/bpb.29.1830.

13. Kostich, W.A., and Sanes, J.R. (1995). Expression of zfh-4, a new member of the zinc finger-homeodomain family, in developing brain and muscle. Developmental Dynamics 202, 145–152. 10.1002/aja.1002020206.

14. Nogami, S., Ishii, Y., Kawaguchi, M., Sakata, N., Oya, T., Takagawa, K., Kanamori, M., Sabit, H., Obata, T., Kimura, T., et al. (2005). ZFH4 protein is expressed in many neurons of developing rat brain. Journal of Comparative Neurology 482, 33–49. 10.1002/cne.20382.

15. Fontana, P., Ginevrino, M., Bejo, K., Cantalupo, G., Ciavarella, M., Lombardi, C., Maioli, M., Scarano, F., Costabile, C., Novelli, A., et al. (2021). *A ZFHX4* mutation associated with a recognizable neuropsychological and facial phenotype. European Journal of Medical Genetics 64, 104321. 10.1016/j.ejmg.2021.104321.

16. Sakata, N., Hemmi, K., Kawaguchi, M., Miura, Y., Noguchi, S., Ma, D., Sasahara, M., Kato, T., Hori, M., and Tamaoki, T. (2000). The Mouse ZFH-4 Protein Contains Four Homeodomains and Twenty-Two Zinc Fingers. Biochemical and Biophysical Research Communications 273, 686–693. 10.1006/bbrc.2000.2990.

17. Nakamura, E., Hata, K., Takahata, Y., Kurosaka, H., Abe, M., Abe, T., Kihara, M., Komori, T., Kobayashi, S., Murakami, T., et al. (2021). Zfhx4 regulates endochondral ossification as the transcriptional platform of Osterix in mice. Commun Biol 4, 1–11. 10.1038/s42003-021-02793-9.

18. Zhang, M., Du, S., Ou, H., Cui, R., Jiang, N., Lin, Y., Ge, R., Ma, D., and Zhang, J. (2021). Ablation of Zfhx4 results in early postnatal lethality by disrupting the respiratory center in mice. J Mol Cell Biol 13, 210–224. 10.1093/jmcb/mjaa081.

19. Chudnovsky, Y., Kim, D., Zheng, S., Whyte, W.A., Bansal, M., Bray, M.-A., Gopal, S., Theisen, M.A., Bilodeau, S., Thiru, P., et al. (2014). ZFHX4 interacts with the NuRD core member CHD4 and regulates the glioblastoma tumor-initiating cell state. Cell Rep 6, 313–324. 10.1016/j.celrep.2013.12.032.

20. Qing, T., Zhu, S., Suo, C., Zhang, L., Zheng, Y., and Shi, L. (2017). Somatic mutations in ZFHX4 gene are associated with poor overall survival of Chinese esophageal squamous cell carcinoma patients. Sci Rep 7, 4951. 10.1038/s41598-017-04221-7.

21. Ha, M., Kim, J., Park, S.M., Hong, C.M., Han, M.-E., Song, P., Kang, C.-D., Lee, D., Kim, Y.H., Hur, J., et al. (2020). Prognostic Role of Zinc Finger Homeobox 4 in Ovarian Serous Cystadenocarcinoma. Genet Test Mol Biomarkers 24, 145–149. 10.1089/gtmb.2019.0185.

22. Zong, S., Xu, P., Xu, Y., and Guo, Y. (2022). A bioinformatics analysis: ZFHX4 is associated with metastasis and poor survival in ovarian cancer. Journal of Ovarian Research 15, 90. 10.1186/s13048-022-01024-x.

23. Millstein, J., Budden, T., Goode, E.L., Anglesio, M.S., Talhouk, A., Intermaggio, M.P., Leong, H.S., Chen, S., Elatre, W., Gilks, B., et al. (2020). Prognostic gene expression signature for high-grade serous ovarian cancer. Ann Oncol 31, 1240–1250. 10.1016/j.annonc.2020.05.019.

24. Sobreira, N., Schiettecatte, F., Valle, D., and Hamosh, A. (2015). GeneMatcher: A Matching Tool for Connecting Investigators with an Interest in the Same Gene. Human Mutation 36, 928–930. 10.1002/humu.22844.

25. Mak, B.C., Sanchez Russo, R., Gambello, M.J., Fleischer, N., Black, E.D., Leslie, E., Murphy, M.M., Mulle, J.G., Bassell, G.J., Aberizk, K., et al. (2021). Craniofacial features of 3q29 deletion syndrome: Application of next-generation phenotyping technology. American Journal of Medical Genetics, Part A 185, 2094–2101. 10.1002/ajmg.a.62227.

26. Pérez Baca, M. del R.P., Jacobs, E.Z., Vantomme, L., Leblanc, P., Bogaert, E., Dheedene, A., Cock, L.D., Haghshenas, S., Foroutan, A., Levy, M.A., et al. (2024). Haploinsufficiency of ZFHX3, encoding a key player in neuronal development, causes syndromic intellectual disability. The American Journal of Human Genetics 0. 10.1016/j.ajhg.2024.01.013.

27. Rao, S.S.P., Huntley, M.H., Durand, N.C., Stamenova, E.K., Bochkov, I.D., Robinson, J.T., Sanborn, A.L., Machol, I., Omer, A.D., Lander, E.S., et al. (2014). A 3D Map of the Human Genome at Kilobase Resolution Reveals Principles of Chromatin Looping. Cell 159, 1665–1680. 10.1016/j.cell.2014.11.021.

28. Durand, N.C., Shamim, M.S., Machol, I., Rao, S.S.P., Huntley, M.H., Lander, E.S., and Aiden, E.L. (2016). Juicer Provides a One-Click System for Analyzing Loop-Resolution Hi-C Experiments. Cell Systems 3, 95–98. 10.1016/j.cels.2016.07.002.

29. Li, H., and Durbin, R. (2009). Fast and accurate short read alignment with Burrows-Wheeler transform. Bioinformatics 25, 1754–1760. 10.1093/bioinformatics/btp324.

30. Melo, U.S., Schöpflin, R., Acuna-Hidalgo, R., Mensah, M.A., Fischer-Zirnsak, B., Holtgrewe, M., Klever, M.-K., Türkmen, S., Heinrich, V., Pluym, I.D., et al. (2020). Hi-C Identifies Complex Genomic Rearrangements and TAD-Shuffling in Developmental Diseases. The American Journal of Human Genetics 106, 872–884. 10.1016/j.ajhg.2020.04.016.

31. Kruse, K., Hug, C.B., and Vaquerizas, J.M. (2020). FAN-C: a feature-rich framework for the analysis and visualisation of chromosome conformation capture data. Genome Biology 21, 303. 10.1186/s13059-020-02215-9.

32. QCloud: A cloud-based quality control system for mass spectrometry-based proteomics laboratories | PLOS ONE https://journals.plos.org/plosone/article?id=10.1371/journal.pone.0189209.

33. Olivella, R., Chiva, C., Serret, M., Mancera, D., Cozzuto, L., Hermoso, A., Borràs, E., Espadas, G., Morales, J., Pastor, O., et al. (2021). QCloud2: An Improved Cloud-based Quality-Control System for Mass-Spectrometry-based Proteomics Laboratories. J. Proteome Res. 20, 2010–2013. 10.1021/acs.jproteome.0c00853.

34. Zhang, X., Smits, A.H., van Tilburg, G.B., Ovaa, H., Huber, W., and Vermeulen, M. (2018). Proteome-wide identification of ubiquitin interactions using UbIA-MS. Nat Protoc 13, 530–550. 10.1038/nprot.2017.147.

35. Ritchie, M.E., Phipson, B., Wu, D., Hu, Y., Law, C.W., Shi, W., and Smyth, G.K. (2015). Limma powers differential expression analyses for RNA-sequencing and microarray studies. Nucleic Acids Research 43, e47. 10.1093/nar/gkv007.

36. Chen, E.Y., Tan, C.M., Kou, Y., Duan, Q., Wang, Z., Meirelles, G.V., Clark, N.R., and Ma’ayan, A. (2013). Enrichr: interactive and collaborative HTML5 gene list enrichment analysis tool. BMC Bioinformatics 14, 128. 10.1186/1471-2105-14-128.

37. Kuleshov, M.V., Jones, M.R., Rouillard, A.D., Fernandez, N.F., Duan, Q., Wang, Z., Koplev, S., Jenkins, S.L., Jagodnik, K.M., Lachmann, A., et al. (2016). Enrichr: a comprehensive gene set enrichment analysis web server 2016 update. Nucleic Acids Research 44, W90–W97. 10.1093/nar/gkw377.

38. Xie, Z., Bailey, A., Kuleshov, M.V., Clarke, D.J.B., Evangelista, J.E., Jenkins, S.L., Lachmann, A., Wojciechowicz, M.L., Kropiwnicki, E., Jagodnik, K.M., et al. (2021). Gene Set Knowledge Discovery with Enrichr. Current Protocols 1, e90. 10.1002/cpz1.90.

39. Lawrence, C., Sanders, G.E., Varga, Z.M., Baumann, D.P., Freeman, A., Baur, B., and Francis, M. (2009). Regulatory Compliance and the Zebrafish. Zebrafish 6, 453–456. 10.1089/zeb.2009.0595.

40. Westerfield, M., Doerry, E., Kirkpatrick, A.E., and Douglas, S.A. (1999). Zebrafish informatics and the ZFIN database. Methods Cell Biol 60, 339–355. 10.1016/s0091-679x(08)61909-3.

41. E3 medium (for zebrafish embryos) (2011). Cold Spring Harb Protoc 2011, pdb.rec66449. 10.1101/pdb.rec066449.

42. Kroll, F., Powell, G.T., Ghosh, M., Gestri, G., Antinucci, P., Hearn, T.J., Tunbak, H., Lim, S., Dennis, H.W., Fernandez, J.M., et al. (2021). A simple and effective F0 knockout method for rapid screening of behaviour and other complex phenotypes. eLife 10, e59683. 10.7554/eLife.59683.

43. CRISPOR: intuitive guide selection for CRISPR/Cas9 genome editing experiments and screens | Nucleic Acids Research | Oxford Academic https://academic.oup.com/nar/article/46/W1/W242/4995687?login=false.

44. inDelphi https://indelphi.giffordlab.mit.edu/.

45. Clement, K., Rees, H., Canver, M.C., Gehrke, J.M., Farouni, R., Hsu, J.Y., Cole, M.A., Liu, D.R., Joung, J.K., Bauer, D.E., et al. (2019). CRISPResso2 provides accurate and rapid genome editing sequence analysis. Nat Biotechnol 37, 224–226. 10.1038/s41587-019-0032-3.

46. Neuhauss, S.C.F., Solnica-Krezel, L., Schier, A.F., Zwartkruis, F., Stemple, D.L., Malicki, J., Abdelilah, S., Stainier, D.Y.R., and Driever, W. (1996). Mutations affecting craniofacial development in zebrafish. Development 123, 357–367. 10.1242/dev.123.1.357.

47. Delbaere, S., Van Damme, T., Syx, D., Symoens, S., Coucke, P., Willaert, A., and Malfait, F. (2020). Hypomorphic zebrafish models mimic the musculoskeletal phenotype of β4GalT7-deficient Ehlers-Danlos syndrome. Matrix Biology 89, 59–75. 10.1016/j.matbio.2019.12.002.

48. Aryee, M.J., Jaffe, A.E., Corrada-Bravo, H., Ladd-Acosta, C., Feinberg, A.P., Hansen, K.D., and Irizarry, R.A. (2014). Minfi: a flexible and comprehensive Bioconductor package for the analysis of Infinium DNA methylation microarrays. Bioinformatics 30, 1363–1369. 10.1093/bioinformatics/btu049.

49. Aref-Eshghi, E., Rodenhiser, D.I., Schenkel, L.C., Lin, H., Skinner, C., Ainsworth, P., Paré, G., Hood, R.L., Bulman, D.E., Kernohan, K.D., et al. (2018). Genomic DNA Methylation Signatures Enable Concurrent Diagnosis and Clinical Genetic Variant Classification in Neurodevelopmental Syndromes. American Journal of Human Genetics 102, 156–174. 10.1016/j.ajhg.2017.12.008.

50. Aref-Eshghi, E., Bend, E.G., Colaiacovo, S., Caudle, M., Chakrabarti, R., Napier, M., Brick, L., Brady, L., Carere, D.A., Levy, M.A., et al. (2019). Diagnostic Utility of Genome-wide DNA Methylation Testing in Genetically Unsolved Individuals with Suspected Hereditary Conditions. The American Journal of Human Genetics 104, 685–700. 10.1016/j.ajhg.2019.03.008.

51. Aref-Eshghi, E., Kerkhof, J., Pedro, V.P., Barat-Houari, M., Ruiz-Pallares, N., Andrau, J.C., Lacombe, D., Van-Gils, J., Fergelot, P., Dubourg, C., et al. (2020). Evaluation of DNA Methylation Episignatures for Diagnosis and Phenotype Correlations in 42 Mendelian Neurodevelopmental Disorders. American Journal of Human Genetics 106, 356–370. 10.1016/j.ajhg.2020.01.019.

52. Ho, D.E., Imai, K., King, G., and Stuart, E.A. (2011). **MatchIt** : Nonparametric Preprocessing for Parametric Causal Inference. J. Stat. Soft. 42. 10.18637/jss.v042.i08.

53. Houseman, E.A., Accomando, W.P., Koestler, D.C., Christensen, B.C., Marsit, C.J., Nelson, H.H., Wiencke, J.K., and Kelsey, K.T. (2012). DNA methylation arrays as surrogate measures of cell mixture distribution. BMC Bioinformatics 13. 10.1186/1471-2105-13-86.

54. Novel diagnostic DNA methylation episignatures expand and refine the epigenetic landscapes of Mendelian disorders - ScienceDirect https://www.sciencedirect.com/science/article/pii/S2666247721000567?ref=cra_js_challenge&fr=RR-1.

55. Ewels, P.A., Peltzer, A., Fillinger, S., Patel, H., Alneberg, J., Wilm, A., Garcia, M.U., Di Tommaso, P., and Nahnsen, S. (2020). The nf-core framework for community-curated bioinformatics pipelines. Nat Biotechnol 38, 276–278. 10.1038/s41587-020-0439-x.

56. Langmead, B., and Salzberg, S.L. (2012). Fast gapped-read alignment with Bowtie 2. Nature Methods 9, 357–359. 10.1038/nmeth.1923.

57. Meers, M.P., Tenenbaum, D., and Henikoff, S. (2019). Peak calling by Sparse Enrichment Analysis for CUT&RUN chromatin profiling. Epigenetics & Chromatin 12, 42. 10.1186/s13072-019-0287-4.

58. Yu, G., Wang, L.-G., and He, Q.-Y. (2015). ChIPseeker: an R/Bioconductor package for ChIP peak annotation, comparison and visualization. Bioinformatics 31, 2382–2383. 10.1093/bioinformatics/btv145.

59. Li, M., Santpere, G., Imamura Kawasawa, Y., Evgrafov, O.V., Gulden, F.O., Pochareddy, S., Sunkin, S.M., Li, Z., Shin, Y., Zhu, Y., et al. (2018). Integrative functional genomic analysis of human brain development and neuropsychiatric risks. Science 362, eaat7615. 10.1126/science.aat7615.

60. Yankee, T.N., Oh, S., Winchester, E.W., Wilderman, A., Robinson, K., Gordon, T., Rosenfeld, J.A., VanOudenhove, J., Scott, D.A., Leslie, E.J., et al. (2023). Integrative analysis of transcriptome dynamics during human craniofacial development identifies candidate disease genes. Nat Commun 14, 4623. 10.1038/s41467-023-40363-1.

61. Uzquiano, A., Kedaigle, A.J., Pigoni, M., Paulsen, B., Adiconis, X., Kim, K., Faits, T., Nagaraja, S., Antón-Bolaños, N., Gerhardinger, C., et al. (2022). Proper acquisition of cell class identity in organoids allows definition of fate specification programs of the human cerebral cortex. Cell 185, 3770–3788.e27. 10.1016/j.cell.2022.09.010.

62. Korsunsky, I., Fan, J., Slowikowski, K., Zhang, F., Wei, K., Baglaenko, Y., Brenner, M., Loh, P.-R., and Raychaudhuri, S. (2018). Fast, sensitive, and accurate integration of single cell data with Harmony. Preprint at bioRxiv, 10.1101/461954.

63. ggplot2.

64. de Sainte Agathe, J.-M., Filser, M., Isidor, B., Besnard, T., Gueguen, P., Perrin, A., Van Goethem, C., Verebi, C., Masingue, M., Rendu, J., et al. (2023). SpliceAI-visual: a free online tool to improve SpliceAI splicing variant interpretation. Human Genomics 17, 7. 10.1186/s40246-023-00451-1.

65. Jaganathan, K., Kyriazopoulou Panagiotopoulou, S., McRae, J.F., Darbandi, S.F., Knowles, D., Li, Y.I., Kosmicki, J.A., Arbelaez, J., Cui, W., Schwartz, G.B., et al. (2019). Predicting Splicing from Primary Sequence with Deep Learning. Cell 176, 535–548.e24. 10.1016/j.cell.2018.12.015.

66. Schenkel, L.C., Aref-Eshghi, E., Rooney, K., Kerkhof, J., Levy, M.A., McConkey, H., Rogers, R.C., Phelan, K., Sarasua, S.M., Jain, L., et al. (2021). DNA methylation epi-signature is associated with two molecularly and phenotypically distinct clinical subtypes of Phelan-McDermid syndrome. Clinical Epigenetics 13, 2. 10.1186/s13148-020-00990-7.

67. Cardoso-Moreira, M., Halbert, J., Valloton, D., Velten, B., Chen, C., Shao, Y., Liechti, A., Ascenção, K., Rummel, C., Ovchinnikova, S., et al. (2019). Gene expression across mammalian organ development. Nature 571, 505. 10.1038/S41586-019-1338-5.

68. Yoshikawa, T., Turner, G., Esterling, L.E., Sanders, A.R., and Detera-Wadleigh, S.D. (1997). A novel human myo-inositol monophosphatase gene, IMP.18p, maps to a susceptibility region for bipolar disorder. Mol Psychiatry 2, 393–397. 10.1038/sj.mp.4000325.

69. Dias, K.-R., Carlston, C.M., Blok, L.E.R., De Hayr, L., Nawaz, U., Evans, C.-A., Bayrak-Toydemir, P., Htun, S., Zhu, Y., Ma, A., et al. (2022). De Novo *ZMYND8* variants result in an autosomal dominant neurodevelopmental disorder with cardiac malformations. Genetics in Medicine 24, 1952–1966. 10.1016/j.gim.2022.06.001.

70. Liu, J., Xiao, Q., Xiao, J., Niu, C., Li, Y., Zhang, X., Zhou, Z., Shu, G., and Yin, G. (2022). Wnt/β-catenin signalling: function, biological mechanisms, and therapeutic opportunities. Sig Transduct Target Ther 7, 1–23. 10.1038/s41392-021-00762-6.

71. Caracci, M.O., Avila, M.E., Espinoza-Cavieres, F.A., López, H.R., Ugarte, G.D., and De Ferrari, G.V. (2021). Wnt/β-Catenin-Dependent Transcription in Autism Spectrum Disorders. Frontiers in Molecular Neuroscience 14, 1–16. 10.3389/fnmol.2021.764756.

72. Carmona, G., Göttig, S., Orlandi, A., Scheele, J., Bäuerle, T., Jugold, M., Kiessling, F., Henschler, R., Zeiher, A.M., Dimmeler, S., et al. (2009). Role of the small GTPase Rap1 for integrin activity regulation in endothelial cells and angiogenesis. Blood 113, 488–497. 10.1182/blood-2008-02-138438.

73. Jaśkiewicz, A., Pająk, B., and Orzechowski, A. (2018). The Many Faces of Rap1 GTPase. International Journal of Molecular Sciences 19, 2848. 10.3390/ijms19102848.

74. Plachez, C., and Richards, L.J. (2005). Mechanisms of Axon Guidance in the Developing Nervous System. In Current Topics in Developmental Biology Neural Development. (Academic Press), pp. 267–346. 10.1016/S0070-2153(05)69010-2.

75. Sarnat, H.B. (2023). Axonal pathfinding during the development of the nervous system. Annals of the Child Neurology Society 1, 13–23. 10.1002/cns3.2.

76. Liu, G., David, B.T., Trawczynski, M., and Fessler, R.G. (2020). Advances in Pluripotent Stem Cells: History, Mechanisms, Technologies, and Applications. Stem Cell Rev and Rep 16, 3–32. 10.1007/s12015-019-09935-x.

77. Swartz, M.E., Sheehan-Rooney, K., Dixon, M.J., and Eberhart, J.K. (2011). Examination of a Palatogenic Gene Program in Zebrafish. Dev Dyn 240, 2204–2220. 10.1002/dvdy.22713.

78. Raterman, S.T., Metz, J.R., Wagener, F.A.D.T.G., and Von den Hoff, J.W. (2020). Zebrafish Models of Craniofacial Malformations: Interactions of Environmental Factors. Front. Cell Dev. Biol. 8. 10.3389/fcell.2020.600926.

79. Pfeiffer, R.A., Hirschfelder, H., and Rott, H.D. (1995). Specific acromesomelia with facial and renal anomalies: a new syndrome. Clin Dysmorphol 4, 38–43.

80. Verloes, A., and David, A. (1995). Dominant mesomelic shortness of stature with acral synostoses, umbilical anomalies, and soft palate agenesis. American Journal of Medical Genetics 55, 205–212. 10.1002/ajmg.1320550211.

81. Isidor, B., Pichon, O., Redon, R., Day-Salvatore, D., Hamel, A., Siwicka, K.A., Bitner-Glindzicz, M., Heymann, D., Kjellén, L., Kraus, C., et al. (2010). Mesomelia-Synostoses Syndrome Results from Deletion of *SULF1* and *SLCO5A1* Genes at 8q13. The American Journal of Human Genetics 87, 95–100. 10.1016/j.ajhg.2010.05.012.

82. Roshandel, D., Sanders, E.J., Shakeshaft, A., Panjwani, N., Lin, F., Collingwood, A., Hall, A., Keenan, K., Deneubourg, C., Mirabella, F., et al. (2023). SLCO5A1 and synaptic assembly genes contribute to impulsivity in juvenile myoclonic epilepsy. npj Genom. Med. 8, 1–11. 10.1038/s41525-023-00370-z.

83. KLF/SP Transcription Factor Family Evolution: Expansion, Diversification, and Innovation in Eukaryotes | Genome Biology and Evolution | Oxford Academic https://academic.oup.com/gbe/article/7/8/2289/557806.

84. ERF: an ETS domain protein with strong transcriptional repressor activity, can suppress ets-associated tumorigenesis and is regulated by phosphorylation during cell cycle and mitogenic stimulation. https://www.embopress.org/doi/epdf/10.1002/j.1460-2075.1995.tb00160.x.

85. Glass, G.E., O’Hara, J., Canham, N., Cilliers, D., Dunaway, D., Fenwick, A.L., Jeelani, N.-O., Johnson, D., Lester, T., Lord, H., et al. (2019). ERF-related craniosynostosis: The phenotypic and developmental profile of a new craniosynostosis syndrome. American Journal of Medical Genetics. Part a 179, 615. 10.1002/ajmg.a.61073.

86. Spruijt, C.G., Luijsterburg, M.S., Menafra, R., Lindeboom, R.G.H., Jansen, P.W.T.C., Edupuganti, R.R., Baltissen, M.P., Wiegant, W.W., Voelker-Albert, M.C., Matarese, F., et al. (2016). ZMYND8 Co-localizes with NuRD on Target Genes and Regulates Poly(ADP-Ribose)-Dependent Recruitment of GATAD2A/NuRD to Sites of DNA Damage. Cell Reports 17, 783–798. 10.1016/j.celrep.2016.09.037.

87. Zhao, D., Guan, H., Zhao, S., Mi, W., Wen, H., Li, Y., Zhao, Y., Allis, C.D., Shi, X., and Li, H. (2016). YEATS2 is a selective histone crotonylation reader. Cell Res 26, 629–632. 10.1038/cr.2016.49.

88. Yeetong, P., Pongpanich, M., Srichomthong, C., Assawapitaksakul, A., Shotelersuk, V., Tantirukdham, N., Chunharas, C., Suphapeetiporn, K., and Shotelersuk, V. (2019). TTTCA repeat insertions in an intron of YEATS2 in benign adult familial myoclonic epilepsy type 4. Brain 142, 3360–3366. 10.1093/brain/awz267.

89. Mi, W., Zhang, Y., Lyu, J., Wang, X., Tong, Q., Peng, D., Xue, Y., Tencer, A.H., Wen, H., Li, W., et al. (2018). The ZZ-type zinc finger of ZZZ3 modulates the ATAC complex-mediated histone acetylation and gene activation. Nat Commun 9, 3759. 10.1038/s41467-018-06247-5.

90. Shinsky, S.A., Monteith, K.E., Viggiano, S., and Cosgrove, M.S. (2015). Biochemical reconstitution and phylogenetic comparison of human SET1 family core complexes involved in histone methylation. J Biol Chem 290, 6361–6375. 10.1074/jbc.M114.627646.

91. Li, N., Li, Y., Lv, J., Zheng, X., Wen, H., Shen, H., Zhu, G., Chen, T.-Y., Dhar, S.S., Kan, P.-Y., et al. (2016). ZMYND8 Reads the Dual Histone Mark H3K4me1-H3K14ac to Antagonize the Expression of Metastasis-Linked Genes. Mol Cell 63, 470–484. 10.1016/j.molcel.2016.06.035.

92. Adhikary, S., Singh, V., Choudhari, R., Yang, B., Adhikari, S., Ramos, E.I., Chaudhuri, S., Roy, S., Gadad, S.S., and Das, C. (2022). ZMYND8 suppresses MAPT213 LncRNA transcription to promote neuronal differentiation. Cell Death Dis 13, 766. 10.1038/s41419-022-05212-x.

93. Lechner, J., Dash, D.P., Muszynska, D., Hosseini, M., Segev, F., George, S., Frazer, D.G., Moore, J.E., Kaye, S.B., Young, T., et al. (2013). Mutational spectrum of the ZEB1 gene in corneal dystrophies supports a genotype-phenotype correlation. Invest Ophthalmol Vis Sci 54, 3215–3223. 10.1167/iovs.13-11781.

94. Williams, T.M., Moolten, D., Burlein, J., Romano, J., Bhaerman, R., Godillot, A., Mellon, M., Rauscher, F.J., and Kant, J.A. (1991). Identification of a zinc finger protein that inhibits IL-2 gene expression. Science 254, 1791–1794. 10.1126/science.1840704.

95. Singh, N.A., Charlier, C., Stauffer, D., DuPont, B.R., Leach, R.J., Melis, R., Ronen, G.M., Bjerre, I., Quattlebaum, T., Murphy, J.V., et al. (1998). A novel potassium channel gene, KCNQ2, is mutated in an inherited epilepsy of newborns. Nat Genet 18, 25–29. 10.1038/ng0198-25.

96. Identification by mass spectrometry and functional characterization of two phosphorylation sites of KCNQ2/KCNQ3 channels 10.1073/pnas.0509122102.

97. Borgatti, R., Zucca, C., Cavallini, A., Ferrario, M., Panzeri, C., Castaldo, P., Soldovieri, M.V., Baschirotto, C., Bresolin, N., Dalla Bernardina, B., et al. (2004). A novel mutation in KCNQ2 associated with BFNC, drug resistant epilepsy, and mental retardation. Neurology 63, 57–65. 10.1212/01.wnl.0000132979.08394.6d.

98. Yang, G., Tian, F., Shen, Y., Yang, C., Yuan, H., Li, P., and Gao, Z. (2023). Functional characterization and in vitro pharmacological rescue of KCNQ2 pore mutations associated with epileptic encephalopathy. Acta Pharmacol Sin 44, 1589–1599. 10.1038/s41401-023-01073-y.

99. Cuvertino, S., Hartill, V., Colyer, A., Garner, T., Nair, N., Al-Gazali, L., Canham, N., Faundes, V., Flinter, F., Hertecant, J., et al. (2020). A restricted spectrum of missense KMT2D variants cause a multiple malformations disorder distinct from Kabuki syndrome. Genet Med 22, 867–877. 10.1038/s41436-019-0743-3.

100. Micale, L., Augello, B., Maffeo, C., Selicorni, A., Zucchetti, F., Fusco, C., De Nittis, P., Pellico, M.T., Mandriani, B., Fischetto, R., et al. (2014). Molecular analysis, pathogenic mechanisms, and readthrough therapy on a large cohort of Kabuki syndrome patients. Hum Mutat 35, 841–850. 10.1002/humu.22547.

101. Shpargel, K.B., Mangini, C.L., Xie, G., Ge, K., and Magnuson, T. (2020). The KMT2D Kabuki syndrome histone methylase controls neural crest cell differentiation and facial morphology. Development 147, dev187997. 10.1242/dev.187997.

102. Sen, R., Lencer, E., Geiger, E.A., Jones, K., Shaikh, T.H., and Artinger, K.B. (2021). The role of Kabuki Syndrome genes KMT2D and KDM6A in development: Analysis in Human sequencing data and compared to mice and zebrafish. Preprint at bioRxiv, 10.1101/2020.04.03.024646.

103. Yoshikawa, T., Padigaru, M., Karkera, J.D., Sharma, M., Berrettini, W.H., Esterling, L.E., and Detera-Wadleigh, S.D. (2000). Genomic structure and novel variants of myo-inositol monophosphatase 2 (IMPA2). Mol Psychiatry 5, 165–171. 10.1038/sj.mp.4000688.

104. Schmidt, A., Marescau, B., Boehm, E.A., Renema, W.K.J., Peco, R., Das, A., Steinfeld, R., Chan, S., Wallis, J., Davidoff, M., et al. (2004). Severely altered guanidino compound levels, disturbed body weight homeostasis and impaired fertility in a mouse model of guanidinoacetate N-methyltransferase (GAMT) deficiency. Hum Mol Genet 13, 905–921. 10.1093/hmg/ddh112.

105. Comeaux, M.S., Wang, J., Wang, G., Kleppe, S., Zhang, V.W., Schmitt, E.S., Craigen, W.J., Renaud, D., Sun, Q., and Wong, L.-J. (2013). Biochemical, molecular, and clinical diagnoses of patients with cerebral creatine deficiency syndromes. Mol Genet Metab 109, 260–268. 10.1016/j.ymgme.2013.04.006.

106. Mercimek-Mahmutoglu, S., Ndika, J., Kanhai, W., de Villemeur, T.B., Cheillan, D., Christensen, E., Dorison, N., Hannig, V., Hendriks, Y., Hofstede, F.C., et al. (2014). Thirteen new patients with guanidinoacetate methyltransferase deficiency and functional characterization of nineteen novel missense variants in the GAMT gene. Hum Mutat 35, 462–469. 10.1002/humu.22511.

107. Nicolas, E., Poitelon, Y., Chouery, E., Salem, N., Levy, N., Mégarbané, A., and Delague, V. (2010). CAMOS, a nonprogressive, autosomal recessive, congenital cerebellar ataxia, is caused by a mutant zinc-finger protein, ZNF592. Eur J Hum Genet 18, 1107–1113. 10.1038/ejhg.2010.82.

108. Hruska-Plochan, M., Wiersma, V.I., Betz, K.M., Mallona, I., Ronchi, S., Maniecka, Z., Hock, E.-M., Tantardini, E., Laferriere, F., Sahadevan, S., et al. (2024). A model of human neural networks reveals NPTX2 pathology in ALS and FTLD. Nature 626, 1073–1083. 10.1038/s41586-024-07042-7.

109. Zhou, J., Wade, S.D., Graykowski, D., Xiao, M.-F., Zhao, B., Giannini, L.A.A., Hanson, J.E., van Swieten, J.C., Sheng, M., Worley, P.F., et al. (2023). The neuronal pentraxin Nptx2 regulates complement activity and restrains microglia-mediated synapse loss in neurodegeneration. Sci Transl Med 15, eadf0141. 10.1126/scitranslmed.adf0141.

110. NPTX2 in Cerebrospinal Fluid Predicts the Progression From Normal Cognition to Mild Cognitive Impairment - Soldan - 2023 - Annals of Neurology - Wiley Online Library https://onlinelibrary.wiley.com/doi/10.1002/ana.26725.

111. Manning, B.J., and Yusufzai, T. (2017). The ATP-dependent chromatin remodeling enzymes CHD6, CHD7, and CHD8 exhibit distinct nucleosome binding and remodeling activities. Journal of Biological Chemistry 292, 11927–11936. 10.1074/jbc.M117.779470.

112. Lutz, T., Stöger, R., and Nieto, A. (2006). CHD6 is a DNA-dependent ATPase and localizes at nuclear sites of mRNA synthesis. FEBS Letters 580, 5851–5857. 10.1016/j.febslet.2006.09.049.

113. Moore, S., Berger, N.D., Luijsterburg, M.S., Piett, C.G., Stanley, F.K.T., Schräder, C.U., Fang, S., Chan, J.A., Schriemer, D.C., Nagel, Z.D., et al. (2019). The CHD6 chromatin remodeler is an oxidative DNA damage response factor. Nat Commun 10, 241. 10.1038/s41467-018-08111-y.

114. Wright, R.H.G., Lioutas, A., Le Dily, F., Soronellas, D., Pohl, A., Bonet, J., Nacht, A.S., Samino, S., Font-Mateu, J., Vicent, G.P., et al. (2016). ADP-ribose–derived nuclear ATP synthesis by NUDIX5 is required for chromatin remodeling. Science 352, 1221–1225. 10.1126/science.aad9335.

115. Yadav, S.P., Sharma, N.K., Liu, C., Dong, L., Li, T., and Swaroop, A. (2016). Centrosomal protein CP110 controls maturation of the mother centriole during cilia biogenesis. Development 143, 1491–1501. 10.1242/dev.130120.

116. Gao, Y., Gan, H., Lou, Z., and Zhang, Z. (2018). Asf1a resolves bivalent chromatin domains for the induction of lineage-specific genes during mouse embryonic stem cell differentiation. Proceedings of the National Academy of Sciences 115, E6162–E6171. 10.1073/pnas.1801909115.

117. Marx, F.P., Holzmann, C., Strauss, K.M., Li, L., Eberhardt, O., Gerhardt, E., Cookson, M.R., Hernandez, D., Farrer, M.J., Kachergus, J., et al. (2003). Identification and functional characterization of a novel R621C mutation in the synphilin-1 gene in Parkinson’s disease. Human Molecular Genetics 12, 1223–1231. 10.1093/hmg/ddg134.

118. Smith, W.W., Liu, Z., Liang, Y., Masuda, N., Swing, D.A., Jenkins, N.A., Copeland, N.G., Troncoso, J.C., Pletnikov, M., Dawson, T.M., et al. (2010). Synphilin-1 attenuates neuronal degeneration in the A53T α-synuclein transgenic mouse model. Human Molecular Genetics 19, 2087–2098. 10.1093/hmg/ddq086.

119. Engelender, S., Kaminsky, Z., Guo, X., Sharp, A.H., Amaravi, R.K., Kleiderlein, J.J., Margolis, R.L., Troncoso, J.C., Lanahan, A.A., Worley, P.F., et al. (1999). Synphilin-1 associates with α-synuclein and promotes the formation of cytosolic inclusions. Nat Genet 22, 110–114. 10.1038/8820.

120. Schoch, K., Meng, L., Szelinger, S., Bearden, D.R., Stray-Pedersen, A., Busk, O.L., Stong, N., Liston, E., Cohn, R.D., Scaglia, F., et al. (2017). A Recurrent De Novo Variant in NACC1 Causes a Syndrome Characterized by Infantile Epilepsy, Cataracts, and Profound Developmental Delay. Am J Hum Genet 100, 343–351. 10.1016/j.ajhg.2016.12.013.

121. Nacc1 Mutation in Mice Models Rare Neurodevelopmental Disorder with Underlying Synaptic Dysfunction | Journal of Neuroscience https://www.jneurosci.org/content/44/14/e1610232024.long.

122. Xie, Q., Tong, C., and Xiong, X. (2024). An overview of the co-transcription factor NACC1: Beyond its pro-tumor effects. Life Sciences 336, 122314. 10.1016/j.lfs.2023.122314.

123. Charlier, C., Singh, N.A., Ryan, S.G., Lewis, T.B., Reus, B.E., Leach, R.J., and Leppert, M. (1998). A pore mutation in a novel KQT-like potassium channel gene in an idiopathic epilepsy family. Nat Genet 18, 53–55. 10.1038/ng0198-53.

124. A Potassium Channel Mutation in Neonatal Human Epilepsy 10.1126/science.279.5349.403.

125. Gómez de San José, N., Massa, F., Halbgebauer, S., Oeckl, P., Steinacker, P., and Otto, M. (2022). Neuronal pentraxins as biomarkers of synaptic activity: from physiological functions to pathological changes in neurodegeneration. J Neural Transm 129, 207–230. 10.1007/s00702-021-02411-2.

126. Antion, M.D., Christie, L.A., Bond, A.M., Dalva, M.B., and Contractor, A. (2010). Ephrin-B3 regulates glutamate receptor signaling at hippocampal synapses. Mol Cell Neurosci 45, 378–388. 10.1016/j.mcn.2010.07.011.

127. Henderson, N.T., Marchand, S.J.L., Hruska, M., Hippenmeyer, S., Luo, L., and Dalva, M.B. (2019). Ephrin-B3 controls excitatory synapse density through cell-cell competition for EphBs. eLife. 10.7554/eLife.41563.

128. Rodenas-Ruano, A., Perez-Pinzon, M.A., Green, E.J., Henkemeyer, M., and Liebl, D.J. (2006). Distinct roles for ephrinB3 in the formation and function of hippocampal synapses. Developmental Biology 292, 34–45. 10.1016/j.ydbio.2006.01.004.

129. Xu, N.-J., Sun, S., Gibson, J.R., and Henkemeyer, M. (2011). A dual-shaping mechanism for postsynaptic ephrin-B3 as a receptor that sculpts dendrites and synapses. Nat Neurosci 14, 1421–1429. 10.1038/nn.2931.

130. Anchoring and synaptic stability of PSD-95 is driven by ephrin-B3 | Nature Neuroscience https://www.nature.com/articles/nn.4140.

131. Rosenbloom, K.R., Armstrong, J., Barber, G.P., Casper, J., Clawson, H., Diekhans, M., Dreszer, T.R., Fujita, P.A., Guruvadoo, L., Haeussler, M., et al. (2015). The UCSC Genome Browser database: 2015 update. Nucleic Acids Research 43, D670--D681. 10.1093/nar/gku1177.

132. Xie, Y., Li, H., Luo, X., Li, H., Gao, Q., Zhang, L., Teng, Y., Zhao, Q., Zuo, Z., and Ren, J. (2022). IBS 2.0: an upgraded illustrator for the visualization of biological sequences. Nucleic Acids Research 50, W420–W426. 10.1093/nar/gkac373.

133. Wilderman, A., VanOudenhove, J., Kron, J., Noonan, J.P., and Cotney, J. (2018). High-Resolution Epigenomic Atlas of Human Embryonic Craniofacial Development. Cell Rep 23, 1581–1597. 10.1016/j.celrep.2018.03.129.

