## Supplemental Figures for "Loss-of-function of the Zinc Finger Homeobox 4 (*ZFHX4*) gene underlies a neurodevelopmental disorder"

### Supplemental Material

#### Supplemental Figures

A.

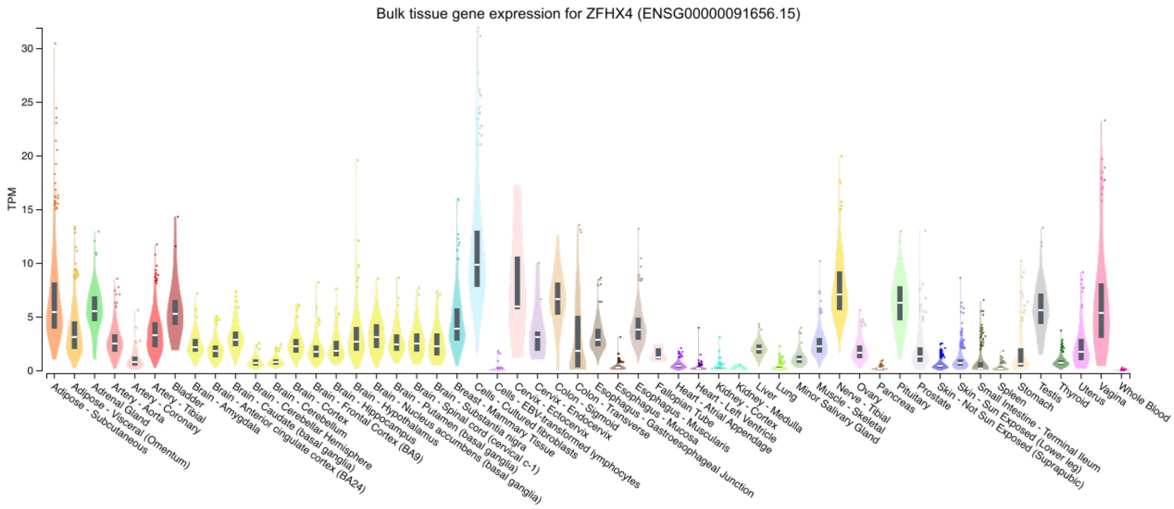


B.

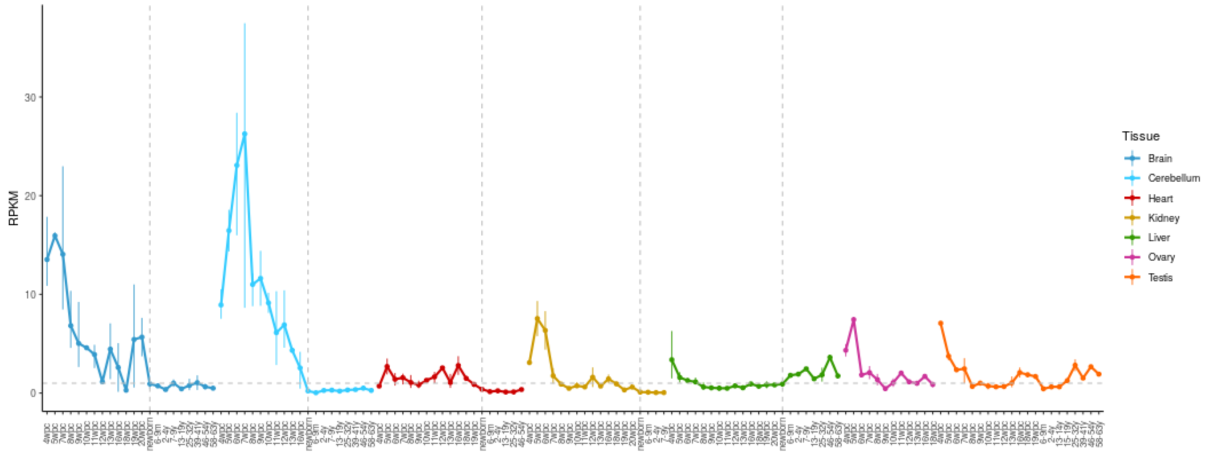


**Figure S1. ZFHX4 expression on the RNA level.** A. in 54 non-diseased adult tissues (GTEx data v8; downloaded from https://gtexportal.org/home/gene/ZFHX4), B. in 22 developmental stages across 7 human tissues (downloaded from https://apps.kaessmannlab.org/evodevoapp/).

A.

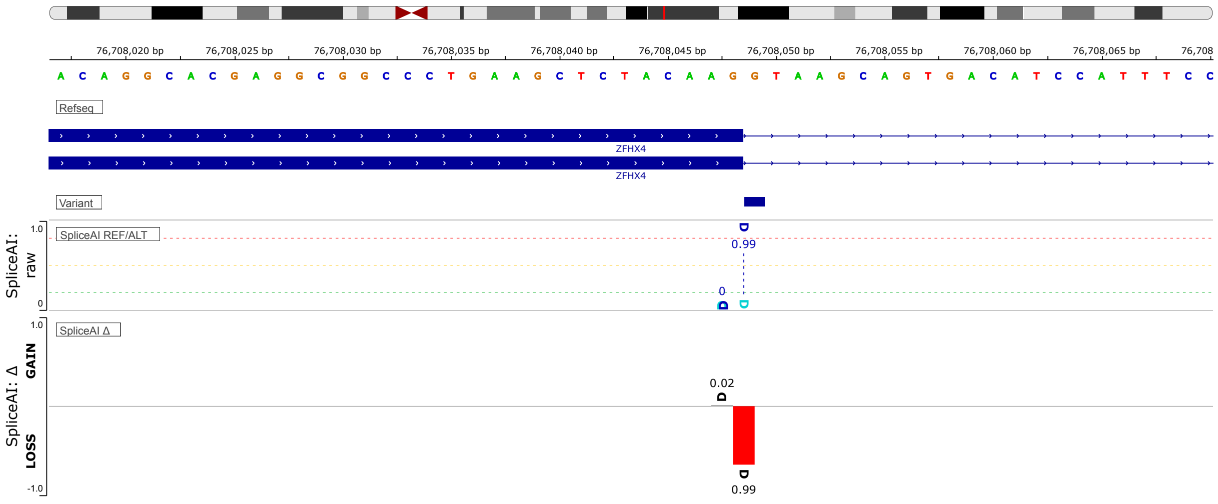


B.

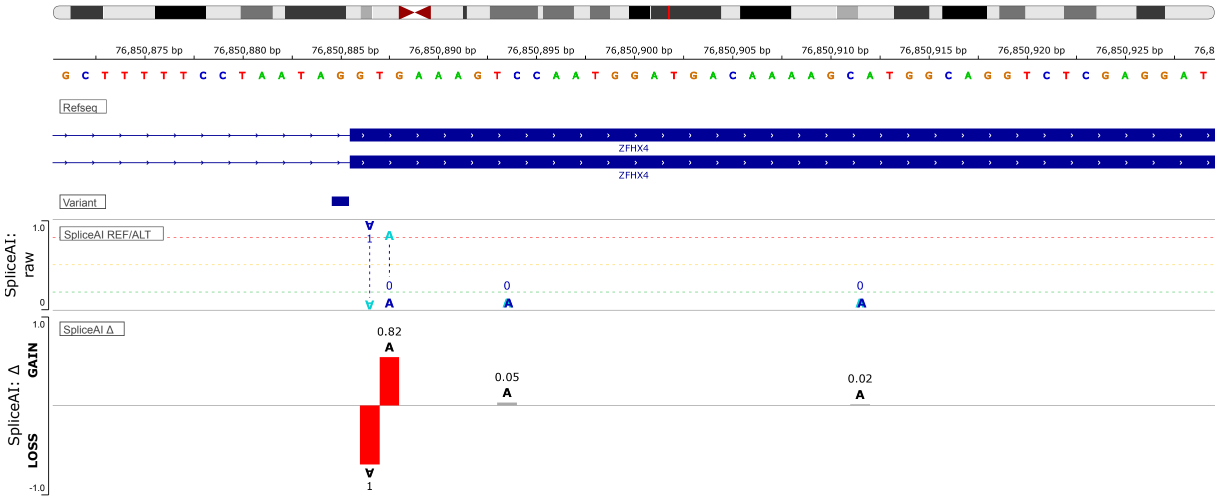


**Figure S2: SpliceAI[1]-visual outputs of ZFHX4 splice variants c.3093+1G>T and c.3965-1G>A displayed in IGV.** A.: SpliceAI raw and delta scores for pathogenic variant (NM_024721.5) ZFHX4 seem to cause the skipping of exon 3 with a stop codon (red). Blue: donor site prediction. The variant position is pointed by a dashed line the variant in individual 33 (ZFHX4 (NM_024721.5: c.3093+1G>T) will result in a donor loss possibly leading to skipping of exon 3. B. SpliceAI raw and delta scores for pathogenic variant (NM_024721.5) ZFHX4 seem to show an acceptor loss followed by acceptor gain (red bar, negative y-axis, positive y-axis). The variant in individual 35 (ZFHX4 (NM_024721.5: c.3965-1G>A) will result in loss of the canonical acceptor at exon 10 and gain of a novel acceptor causing a frameshift on exon 10.

A.

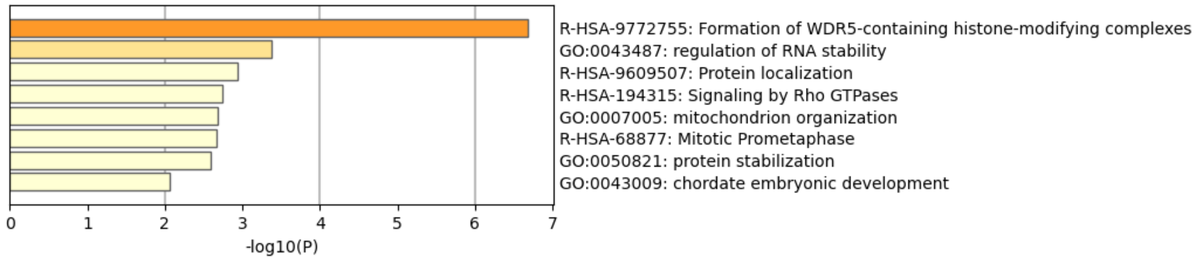


B.

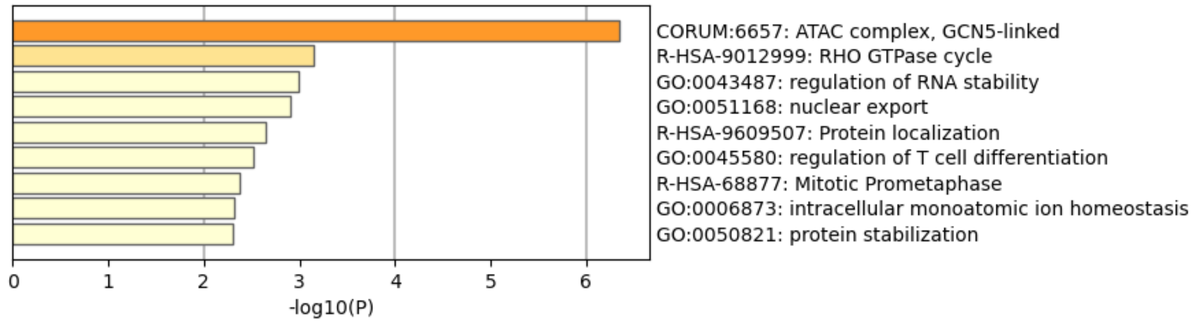


**Figure S3: Metascape[2] enrichment of the ZFHX4 interactome.** Enrichment heatmap of 36 (FDR<0.01) A. and 46 (FDR<0.05) B. enriched GO-terms across input protein lists (= ZFHX4 and 36 or 46 interactors), colored by values.

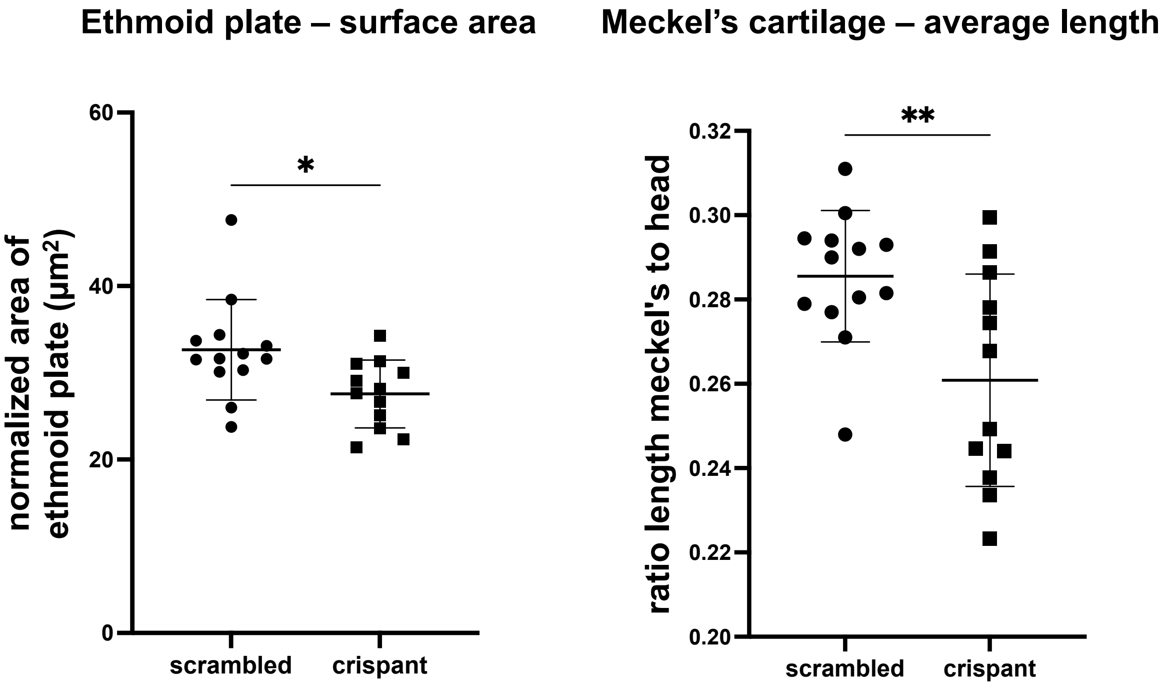


**Figure S4: Zebrafish zfhx4 crispants larvae showed smaller ethmoid plates and shorter Meckel's cartilage structures. Left:** the surface of the ethmoid plate was measured and normalized to the head length of the larvae. **Right:** the length of the Meckel's cartilages was measured and normalized to the head length of the larvae. The Meckel's and the ethmoid plate showed significant differences in the crispant larvae in comparison with the scrambled larvae (p-value ≤ 0.05 (*), p-value ≤ 0.01 (**)).

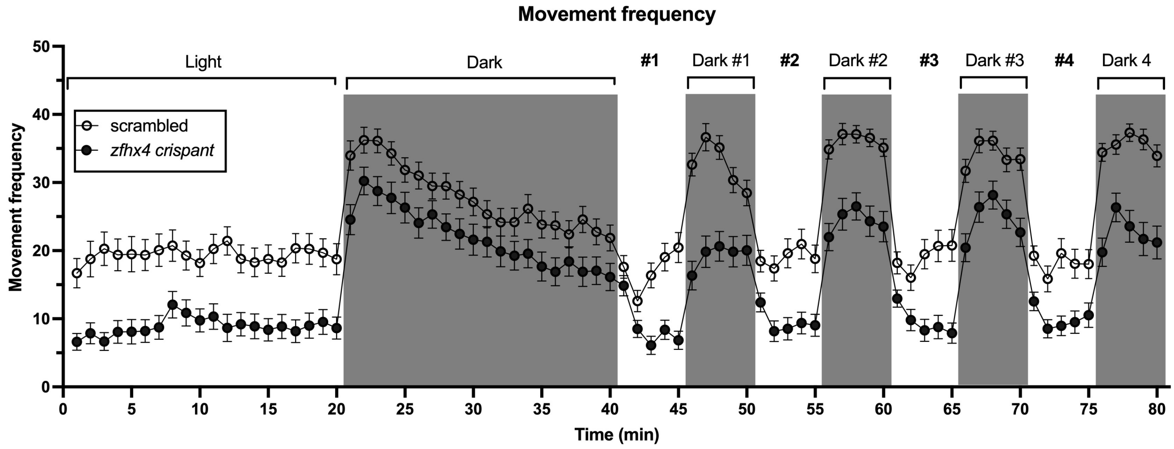


**Figure S5: *zfhx4* crispants show a consistently lower movement frequency compared to scrambled control larvae during the complete behavior experiment but the trends during light and dark cycles remain similar in another experimental replicate**. Dark periods are indicated with grey boxes. Data is represented as mean ± SEM (replicate 1 (above) n = 48/genotype, replicate 2 (bottom) n=60/genotype).

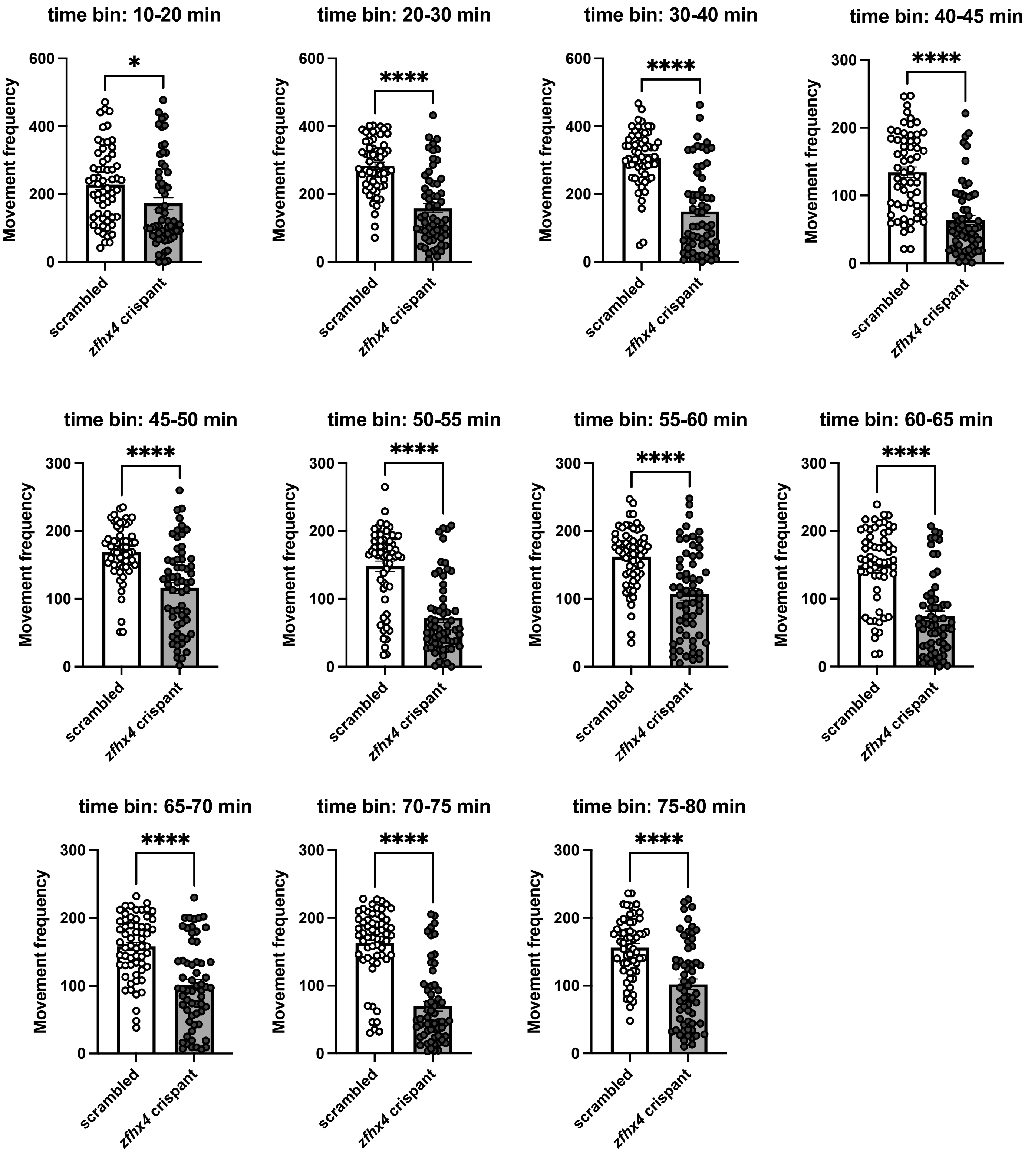


**Figure S6: *zfhx4* crispants show a significantly lower movement frequency compared to scrambled control larvae during the light/dark cycle.** The scrambled and *zfhx4* crispants movement frequency is compared in tile periods. Each period of 10 minutes until the total of 40 minutes, followed by periods of 5 minutes until the total of 80 minutes; are represented in a bar plot with the p-values (p-value ≤ 0.05 (*), p-value ≤ 0.0001 (****)), showing the significant differences in movement frequency between conditions. Data is represented as mean ± SEM (n=60/genotype).

#### References


1. de Sainte Agathe, J.-M., Filser, M., Isidor, B., Besnard, T., Gueguen, P., Perrin, A., Van Goethem, C., Verebi, C., Masingue, M., Rendu, J., et al. (2023). SpliceAI-visual: a free online tool to improve SpliceAI splicing variant interpretation. Human Genomics *17*, 7. https://doi.org/10.1186/s40246-023-00451-1.

2. Zhou, Y., Zhou, B., Pache, L., Chang, M., Khodabakhshi, A.H., Tanaseichuk, O., Benner, C., and Chanda, S.K. (2019). Metascape provides a biologist-oriented resource for the analysis of systems-level datasets. Nature Communications *10*. https://doi.org/10.1038/s41467-019-09234-6.
